# Repurposed antiviral medicines for potential pandemic viruses: A horizon scan

**DOI:** 10.1101/2025.09.09.25335403

**Authors:** Sola Akinbolade, Rhiannon Potter, Alex Inskip, Jane Nesworthy, Kirsti Brock, Gill Norman

## Abstract

**Background:** Viruses such as Ebola, Marburg, influenza, mpox, MERS-CoV, SARS-CoV, and SARS-CoV-2 pose a significant risk for future pandemics. Developing novel antiviral medicines can be time-consuming and resource intensive. Repurposing existing medicines with antiviral activity offers a faster, cost-effective strategy to expand treatment options during public health emergencies. This scan aimed to identify and synthesise recent evidence on repurposed antiviral medicines under investigation for these viruses.

**Method:** A horizon scanning approach was employed, starting with a targeted search in Embase, followed by a systematic search of ClinicalTrials.gov to capture the developmental stages of the technologies. Eligible technologies included UK- or EU-licensed medicines repurposed as antiviral therapies for the viruses of interest. Vaccines, unlicensed medicines, and already approved treatments for the targeted viruses were excluded.

**Results:** A total of 196 repurposed technologies targeting the viruses were identified from published literature, and the expanded search on the clinical trials registry yielded 58 technologies in active clinical development. Interventional clinical trial activity was limited to influenza and COVID-19, with 29 technologies for COVID-19 and two for influenza advancing to phase III evaluation. For other viruses, proposed antiviral candidates were identified in the literature but had not progressed into clinical development. Commonly investigated pharmacological classes included direct-acting antivirals, tyrosine kinase inhibitors, immunomodulators, and anti-inflammatory agents.

**Conclusion:** Repurposing antiviral medicines represents a pragmatic strategy for rapid therapeutic deployment against emerging viral threats. Collaboration among researchers, policymakers, research funders, and regulatory bodies will be essential to improve pandemic preparedness and support repurposing efforts in emergency situations.

## Introduction

Viruses are microscopic organisms capable of infecting hosts, though not all are contagious. Those that are transmissible can spread rapidly within human populations, and some are considered potential pandemic threats due to their ability to cause significant illness and/or death.^1^ A virus cannot replicate outside of living cells of a host organism, however, once it infiltrates a host cell, it uses components within the host cell to multiply, often killing the host cell and causing damage to the host organism.^2^ Viruses contain either a RNA (ribonucleic acid) or DNA (deoxyribonucleic acid) genome surrounded by a virus-coded protein coat.^2^ Genetic changes or mutations in these genetic materials result in new functions or enhanced characteristics of the virus such as its ability to cause different variants of the viral disease.^1^

Pandemics caused by viruses may originate from either zoonotic (animal-to-human) or non-zoonotic (human-to-human or environmental) transmission pathways. For instance, Ebola and Marburg viruses are zoonotic RNA filoviruses typically transmitted through direct contact with infected bodily fluids. These viruses are believed to have been contracted from wild animals and then spread to humans, followed by human-to-human transmission.^3^ Influenza virus, another potential pandemic virus, is a highly contagious respiratory virus responsible for annual global epidemics of seasonal infections. Periodic emergence of new influenza strains – against which there is little or no pre-existing immunity – can lead to widespread severe illness, particularly in individuals with underlying health conditions.^4^ Smallpox and mpox (formerly known as monkeypox) are infectious diseases caused by viruses belonging to the Orthopoxvirus family.^5,6^ Mpox can be transmitted zoonotically, typically through bites or scratches from infected animals.^6^ Notably, smallpox was eradicated in the 1970s through a successful global vaccination programme.^7^ Other notable zoonotic pandemic threats include MERS-CoV (Middle East respiratory syndrome coronavirus), SARS-CoV (severe acute respiratory syndrome coronavirus), and SARS-CoV-2, which are RNA coronaviruses capable of airborne transmission and rapid human-to-human spread.^3^

Historically, viral pandemics have caused high mortality and continue to pose a global threat due to emerging and re-emerging outbreaks, driven by evolution of viral variants.^3^ These challenges highlight the critical importance of having effective therapeutic options readily available for future pandemic responses. Promising strategies for combating viral infections often involve direct-acting antivirals which target specific components of the virus, or host-targeted therapies that modulate the host immune response to inhibit viral replication and minimise immune-related damage.^8^

The discovery and development of novel medicines often face significant challenges, including long timelines, high costs, and the risk of unforeseen adverse events. These are further compounded by complex regulatory hurdles, which can increase expenses and delay the introduction of new therapies to market.^9,10^ Existing antiviral medicines originally developed and approved for specific infections can be evaluated for their efficacy and effectiveness against other viral threats. This strategy, known as medicine repurposing or repositioning, involves identifying new therapeutic uses for already approved medicines outside the scope of their original indication.^9,10^ Repurposed medicines often have established safety profiles, which can reduce the associated risk of novel drug development.^9^ There is potentially a reduction in both development time and cost, largely due to being able to use existing data from completed preclinical studies, safety assessments, and, in some cases, formulation development.^10^ As a result, repurposing efforts can often begin from proof-of-concept phase II clinical trials, with the primary objectives of evaluating efficacy, as well as assessing safety and tolerability.^11^ The reduction in development time and cost associated with repurposing can be particularly valuable in a viral pandemic where immediate treatment options are crucial for controlling the spread and severity of the disease. Repurposing existing direct-acting antivirals or host-targeted agents could offer a potential pathway for developing effective treatments against various viral diseases.

This scan aims to explore the pipeline of repurposed medicines with known or potential antiviral activity against potential pandemic viruses, including Ebola, Marburg, influenza, mpox, smallpox, MERS-CoV, SARS-CoV, and SARS-CoV-2. Additionally, it seeks to offer insights and strategic guidance to support future antiviral repurposing efforts.

## Method

A horizon scanning approach was employed to identify antiviral medicines currently in development or under investigation for potential repurposing. This strategy involved systematically searching bibliographic and clinical trials databases to capture emerging innovations and potential therapeutic opportunities within the antiviral field.^12^

The scan for antiviral medicines encompassed those traditionally classified as ‘conventional’ or direct-acting antivirals as well as medicines from other therapeutic classes that possess known or potential antiviral activity, which may be repurposed for the treatment of emerging or re-emerging pandemic viruses.

### Eligibility criteria

To ensure a systematic and transparent approach, predefined inclusion/exclusion criteria were established to determine the eligibility of studies, as follows (Table 1):

**Table 1.**
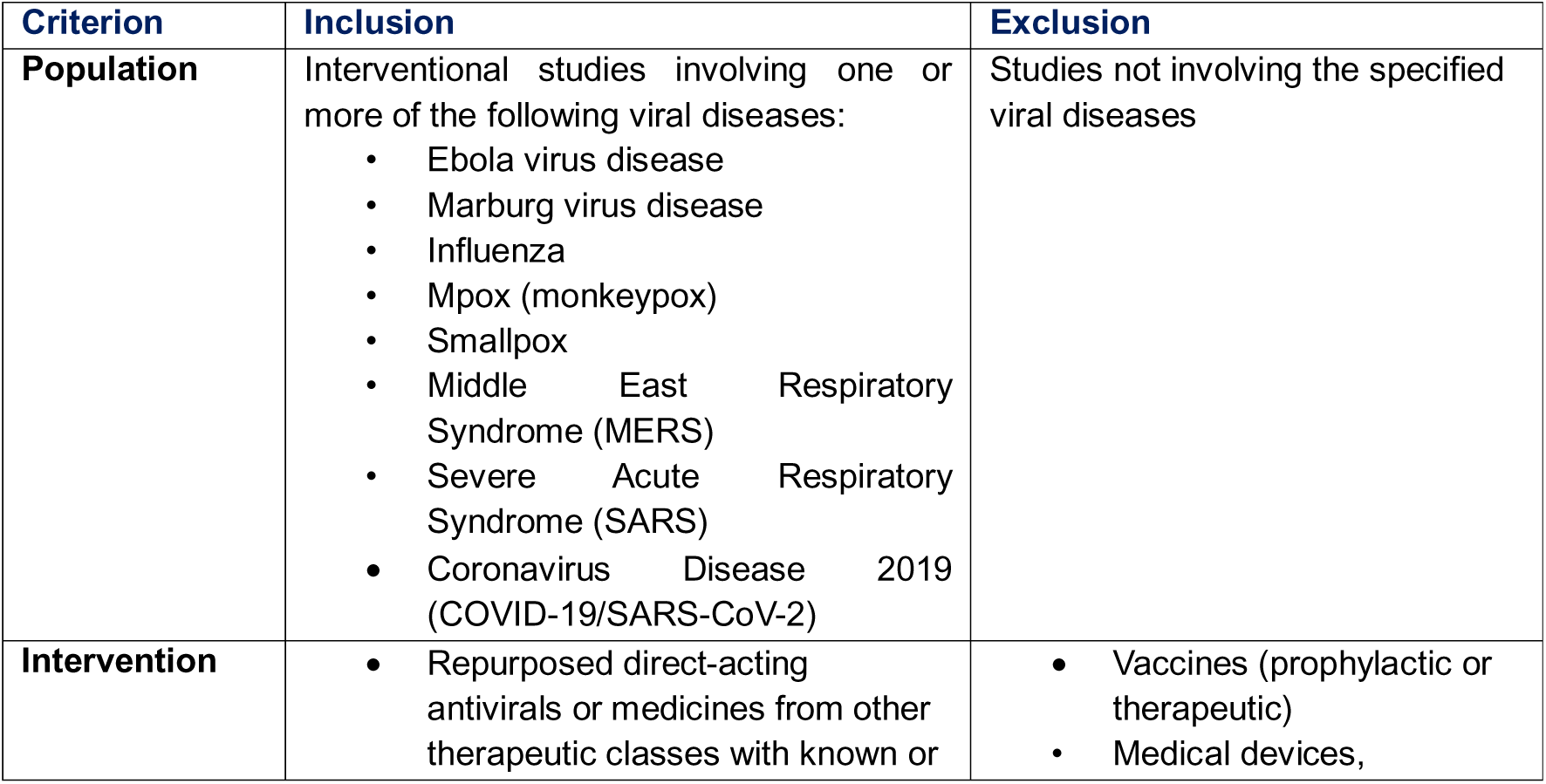
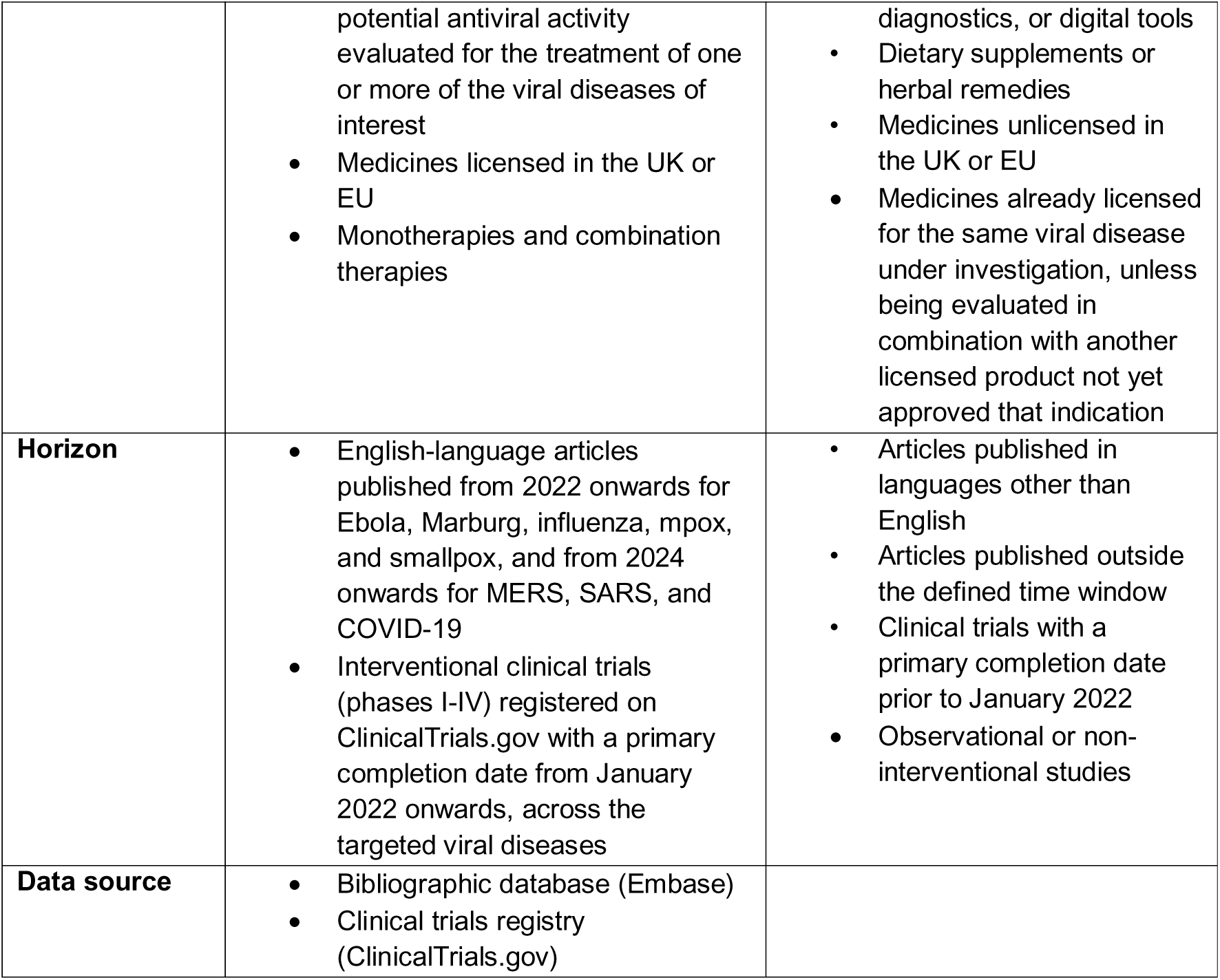
Eligibility criteria.

### Search Strategy

#### Literature search

A literature search was conducted in April 2025 using the Embase^13^ bibliographic database to identify recent publications relevant to the development of repurposed antiviral medicines. The search was limited to articles published from 2022 onwards for Ebola, Marburg, influenza, mpox, and smallpox. For MERS, SARS, and COVID-19, the search was restricted to publications from 2024 onwards to focus on more refined, post-pandemic strategic research and to limit the volume of data.

The search strategy incorporated a broad range of keywords, including terms such as ‘antivirus agent’, ‘antiviral therapy’, ‘antiviral activity’, ‘drug repurposing’, ‘drug repositioning’, ‘off-label drug use’, ‘licensed’, ‘MHRA-approved’, ‘EU-approved’, ‘UK-approved’, as well as disease-specific terms and synonyms for each target virus. The full search strategy, including all search strings and filters, is provided in Appendix A.

All identified articles were exported into Microsoft Excel for manual screening against the predefined inclusion criteria, followed by extraction of eligible medicinal products. This process ensured a systematic and consistent identification of relevant medicinal products.

#### Clinical trials search

Following the identification of technologies from the literature search, their known aliases and alternative names were systematically used to search the ClinicalTrials.gov^14^ registry for interventional studies, accounting for variability in how products may be reported. This approach ensured comprehensive capture of their developmental stages.

To further broaden the search scope, additional searches were conducted without specifying product names, focusing instead on trial records containing the term ‘antiviral’ (or ‘anti-viral’) in the title or summary. This multi-layered strategy maximised the identification of relevant antiviral medicinal products while maintaining a manageable volume of results.

To focus on the most recent therapeutic developments, only trials with a primary completion date from January 2022 onwards were included. All eligible records were exported to Microsoft Excel for manual screening and further assessment.

### Study screening and selection

All identified studies were independently screened in Microsoft Excel against the predefined eligibility criteria by members of the review team (SA, RP, JN, or KB). Each study was assessed and marked as either ‘include’ or ‘exclude’ based on its relevance to the study objectives. If a screener encountered uncertainty or had a query about a particular study, it was flagged for further review. These flagged studies were then assessed by a second reviewer, and any discrepancies were resolved through discussion to reach consensus on inclusion or exclusion. This screening process aimed to enhance consistency, reduce bias, and ensure rigorous selection of relevant studies.

### Data Extraction

Data were manually extracted from all included studies to generate an overview of each identified antiviral medicine, detailing the medicine name (whether investigated as a monotherapy or in combination) and its corresponding targeted viral disease. Extraction was conducted using a standardised Microsoft Excel data extraction form to ensure consistency across reviewers. Any uncertainties or discrepancies encountered during the extraction process were documented and resolved through discussion within the full review team. Upon completion of data extraction, a quality assurance check was performed by a second reviewer to ensure accuracy and consistency of the extracted information.

### Verification of repurposing status

To verify regulatory approval status of each identified antiviral medicine, the UK and EU licensing information was manually cross-referenced using the electronic medicines compendium^15^ or European Medicines Agency^16^ websites respectively. This step ensured that all shortlisted medicines were already approved for use in at least one indication by either the UK or EU regulatory authorities, and that unlicensed medicines were excluded. Additionally, this verification process helped identify whether each medicine had already been licensed for the treatment of any of the targeted viral diseases – Ebola, Marburg, influenza, mpox, smallpox, MERS, SARS, or COVID-19. Medicines already licensed for a specific viral disease were excluded if they were being evaluated for the same viral disease indication. However, an exception was made for medicines already licensed for a specific viral disease if they were being evaluated in combination with another licensed medicine that had not yet been approved for that viral indication. This approach allowed the review to capture novel combination therapies with potential to advance antiviral treatment options beyond current standards of care.

## Results

The search of the bibliographic database and the subsequent search of the clinical trials registry yielded 274 and 371 results, respectively (Figure 1). After thorough screening against the predefined eligibility criteria, 112 articles and 30 clinical trial records were included in the analysis. The medicinal products identified were classified as ‘technologies’, which we define as either a single medicine or a combination of medicines investigated for the treatment of a specific viral indication.

**Figure 1.**
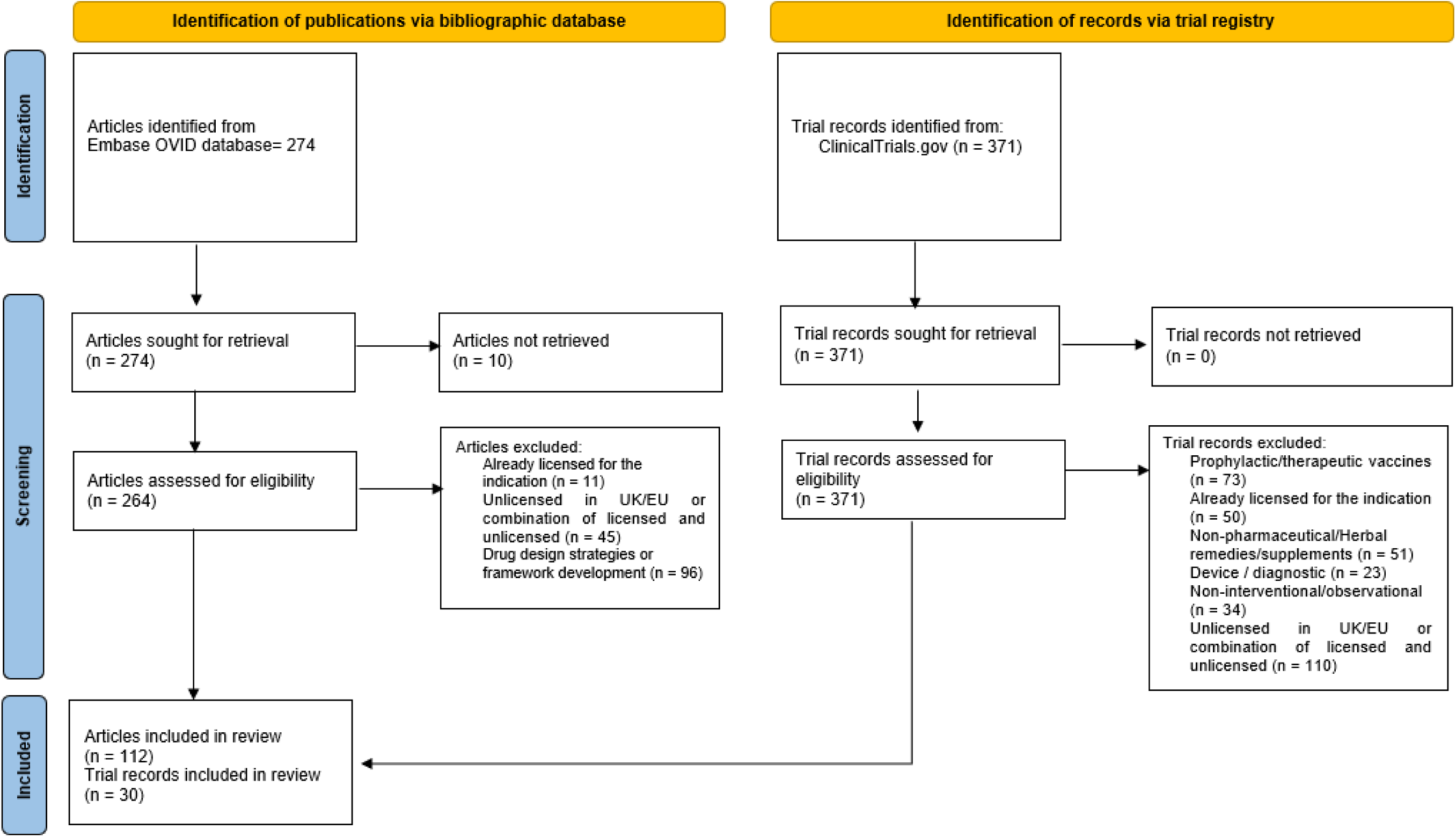
Flow diagram illustrating the selection process and the final number of included articles and clinical trial records

From the 112 articles identified through the literature search, 196 unique technologies were extracted. In parallel, the 30 clinical trial records yielded 58 technologies, as several trials investigated multiple therapeutic interventions across different treatment arms. This reflects the wide range of treatment strategies being explored within individual trials. Some of the technologies involved medicines already licensed for one of the target viral diseases but being investigated in combination with another licensed medicine not yet approved for that specific viral indication, thus representing novel therapeutic approaches.

Figure 1 presents the flow diagram^17^ illustrating the process of study identification, screening, and inclusion.

Repurposed antiviral technologies were identified from published literature for Ebola, Marburg, influenza, mpox, MERS, SARS, and COVID-19. No eligible technologies were identified for smallpox, consistent with its successful eradication in the late 20th century through global vaccination programmes. The identified technologies were either in preclinical development, under clinical investigation or proposed as potential candidates for future repurposing initiatives. They are grouped by their target viral disease (Figure 2) and are described in detail in the following sections.

**Figure 2.**
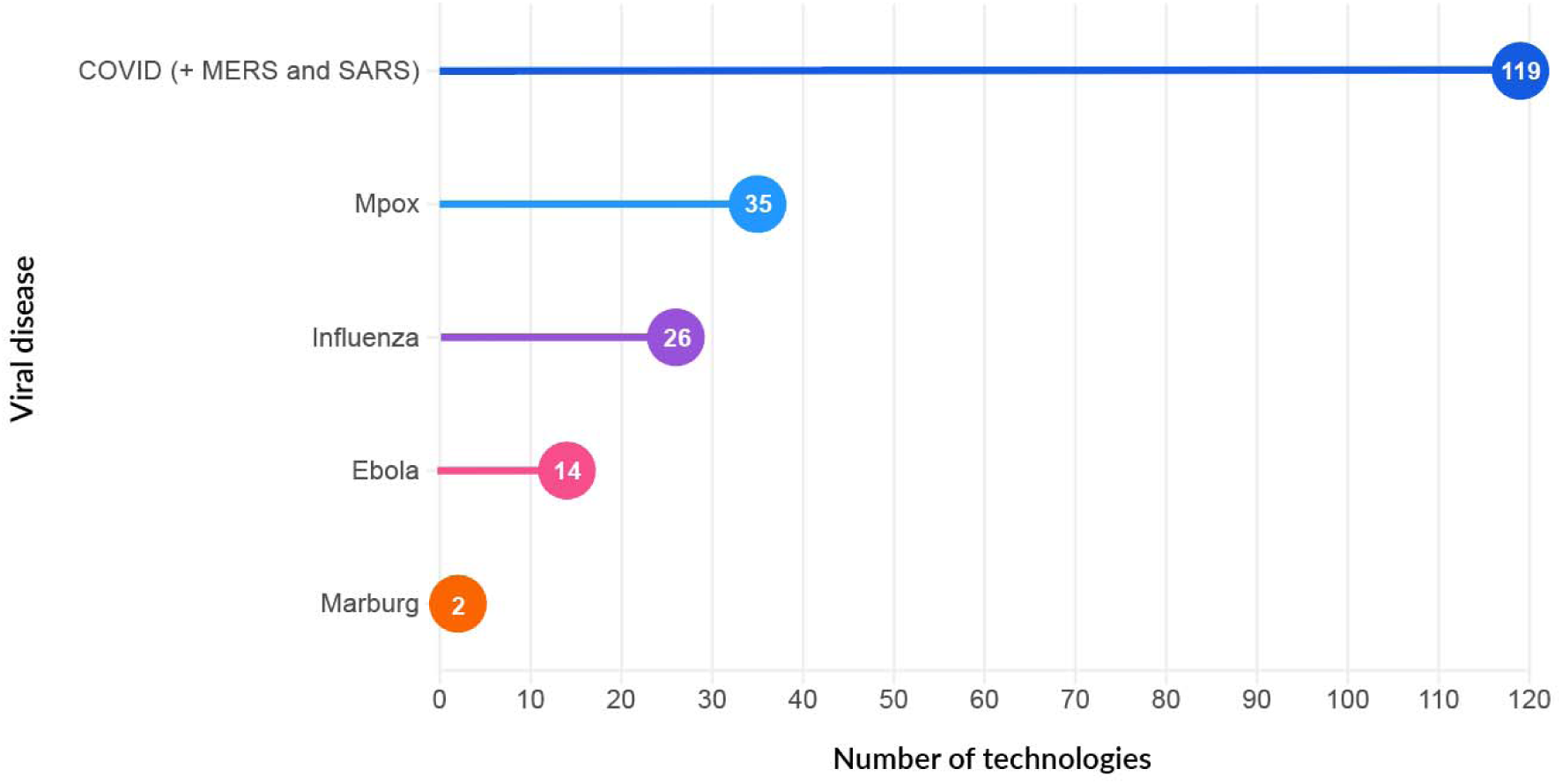
Overview of identified technologies from published literature

### Ebola

The analysis of published literature identified fourteen technologies proposed for repurposing as potential treatments for Ebola virus disease (EVD). All were originally approved for indications other than Ebola. Among them, only three were direct-acting antivirals; the rest included diverse classes such as selective oestrogen receptor modulators, antibiotics, and thrombopoietin receptor agonists. Notably, two of these repurposed candidates were originally approved as cancer therapies (see Supplementary Table 1).

EVD is characterised by the virus’s ability to target and destroy immune cells, leading to immune suppression and the onset of severe disease.^18^ The progression and outcome of EVD depend largely on the host’s ability to suppress viral replication and the virus’s ability to evade or manipulate the immune response.^18^ As such, treatment strategies identified in literature for EVD focused on approaches such as the use of direct-acting antivirals targeting the virus itself (e.g. glecaprevir, velpatasvir), agents that block viral entry or replication (e.g., selective oestrogen receptor modulators [tamoxifen, raloxifene]), therapies that modulate host immune responses (e.g., selective serotonin reuptake inhibitors [fluoxetine]), or statins for their anti-inflammatory effect and vascular protection (e.g. simvastatin) (Figure 3) (full details available in Supplementary Table 1).

**Figure 3.**
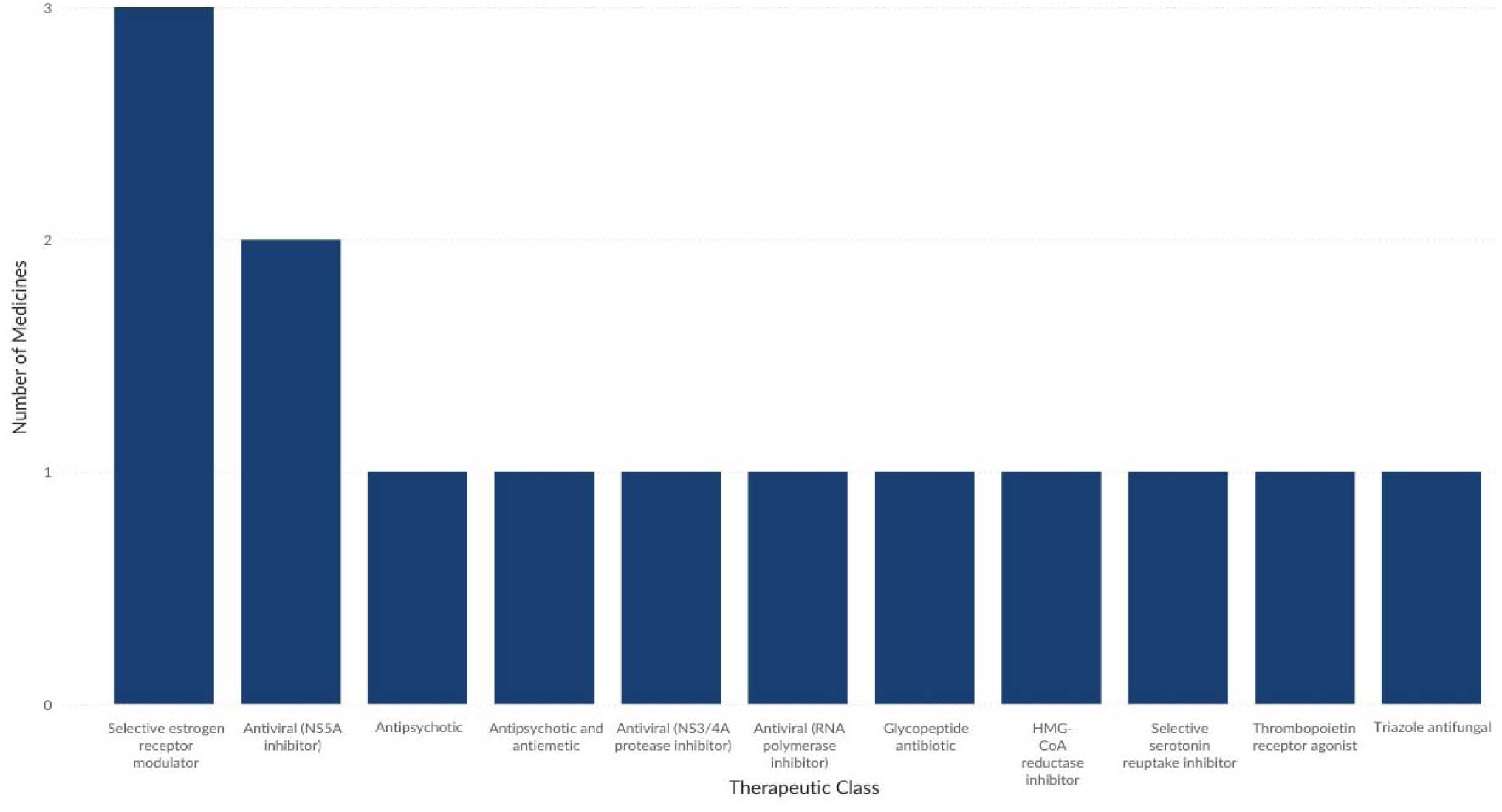
Number of identified technologies per therapeutic class evaluating treatments for Ebola virus disease

### Marburg

According to the World Health Organisation, there are currently no approved antiviral therapies for the treatment of Marburg virus disease, and potential candidates remain under investigation.^19^ In the absence of licensed therapeutic options, current development efforts appear to prioritise the evaluation of existing antivirals for potential repurposing. Our analysis identified two candidates under investigation for Marburg virus disease, namely remdesivir, an RNA polymerase inhibitor originally approved for the treatment of COVID-19, and bictegravir, an integrase strand transfer inhibitor used in the management of human immunodeficiency virus infection. Both medicines have demonstrated antiviral activity in their original indications, supporting their exploration as potential therapies against Marburg virus disease (full details available in Supplementary Table 2).

### Influenza

Antiviral medicines approved for the treatment of influenza have demonstrated the ability to reduce symptom severity and lower the risk of complications when administered early in the course of infection.^20^ Ongoing research is increasingly focused on developing therapeutic options that may effectively combat the virus while mitigating the emergence of drug-resistant viral strains.^21^

Our analysis identified 26 technologies currently being investigated for potential repurposing in the treatment of influenza virus infection. These candidates represent a broad spectrum of therapeutic classes, including direct-acting antivirals (e.g. molnupiravir); imidazole antifungals (e.g. econazole) with potential host-directed antiviral effects; thrombopoietin receptor agonists (e.g. eltrombopag) aimed at enhancing platelet production; tyrosine kinase inhibitors (e.g. afatinib) that may interfere with viral entry and replication; and antineoplastic agents (e.g. idarubicin) with immunomodulatory properties (Figure 4) (full details available in Supplementary Table 3). All identified candidates were originally developed and approved for indications unrelated to influenza, encompassing both cancer and non-cancer conditions, highlighting the potential of cross-indication repurposing strategies.

**Figure 4.**
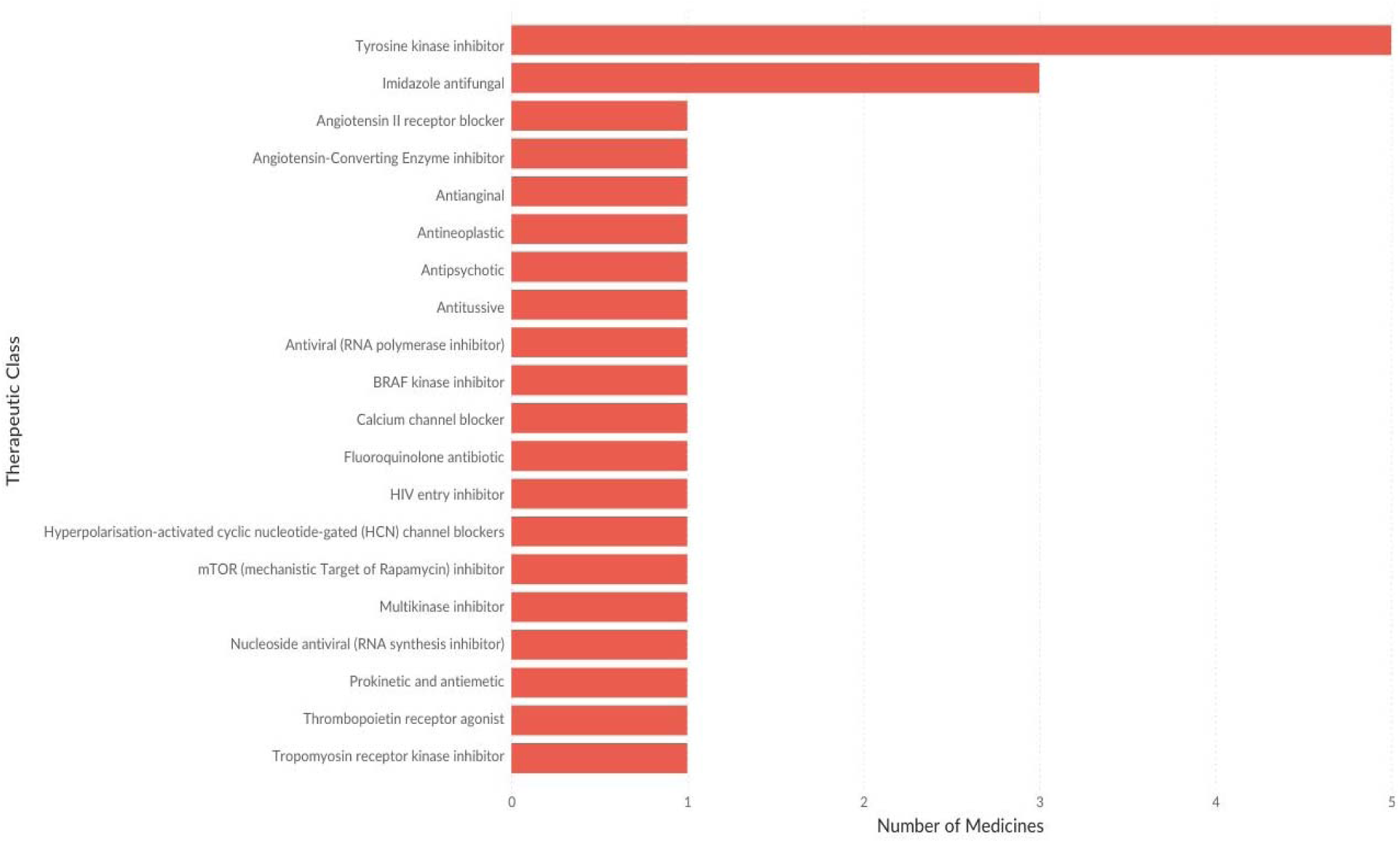
Number of identified repurposing technologies per therapeutic class for influenza treatment

### Mpox

Thirty-five technologies were identified in published literature as candidates for repurposing in the treatment of mpox virus. Among these, two were combination therapies involving tecovirimat (currently the only approved antiviral for mpox)^22,23^ paired with medicines licensed for non-mpox indications. The first combination included fenofibrate, a lipid-lowering agent approved for the treatment of hypertriglyceridaemia and mixed hyperlipidaemia, administered alongside tecovirimat. The second involved mycophenolate, an immunosuppressive agent approved for the prophylaxis of acute transplant rejection, also combined with tecovirimat. The exclusive approval of tecovirimat for the treatment of mpox highlights the critical need to explore additional treatment options, especially in cases of emerging drug resistance, hypersensitivity reactions, or inadequate clinical response.^24^

The remaining identified technologies were monotherapies exploring the repurposing potential of diverse pharmacological classes. These included antivirals (e.g. baloxavir, cidofovir) aimed at directly inhibiting viral replication; integrase strand transfer inhibitors (e.g. cabotegravir, dolutegravir) investigated for their potential host-modulatory effects; tumour necrosis factor inhibitors (e.g. adalimumab, infliximab) used for their immunosuppressive properties to mitigate inflammation; and tyrosine kinase inhibitors (e.g. ponatinib, nilotinib) which may disrupt viral entry and replication (Figure 5) (full details available in Supplementary Table 4). This breadth of therapeutic classes reflects a growing interest in host-targeted and multi-mechanism approaches for the treatment of mpox.

**Figure 5.**
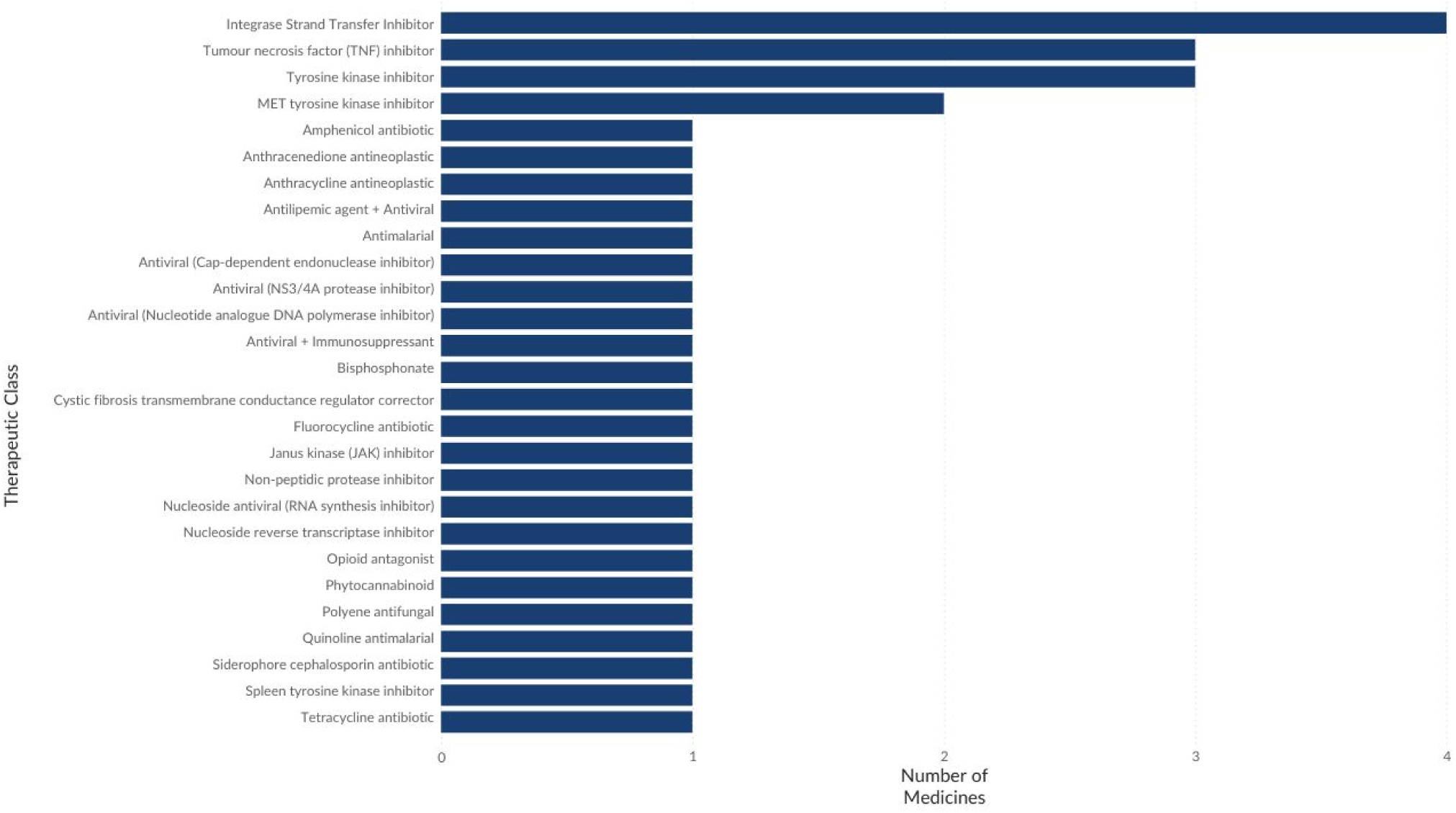
Number of identified repurposing technologies per therapeutic class for mpox treatment

### MERS, SARS, and COVID-19

Middle East respiratory syndrome coronavirus (MERS-CoV), severe acute respiratory syndrome coronavirus (SARS-CoV), and SARS-CoV-2 are closely related RNA viruses responsible for outbreaks of severe respiratory disease with high case fatality rates. These coronaviruses exhibit considerable overlap in clinical presentation, including fever, cough, and respiratory distress, making them clinically difficult to distinguish based on symptoms alone.^25^

A total of 119 technologies were identified from literature as potential repurposing options for the treatment of coronavirus infections. Of these, only nine were proposed for use across MERS, SARS, and COVID-19, while the remainder were exclusively evaluated for COVID-19. Among the 119 candidates, 21 were originally approved for oncological indications, whereas the rest were developed for non-cancer conditions, reflecting a broad therapeutic base for repurposing efforts.

These technologies encompassed a wide range of pharmacological classes, including direct-acting antivirals such as zanamivir and ledipasvir, which target viral replication pathways. Antimalarial agents like chloroquine diphosphate and the combination of pyronaridine with artesunate were explored for their potential antiviral and immunomodulatory properties. Tyrosine kinase inhibitors, including entrectinib and nilotinib, were investigated for their ability to disrupt viral entry and replication. Selective serotonin reuptake inhibitors such as fluoxetine and fluvoxamine were evaluated for their immunomodulatory effects, while alkylating agents like carboplatin and cisplatin were considered for their cytotoxic activities (Figure 6) (full details available in Supplementary Table 5).

**Figure 6.**
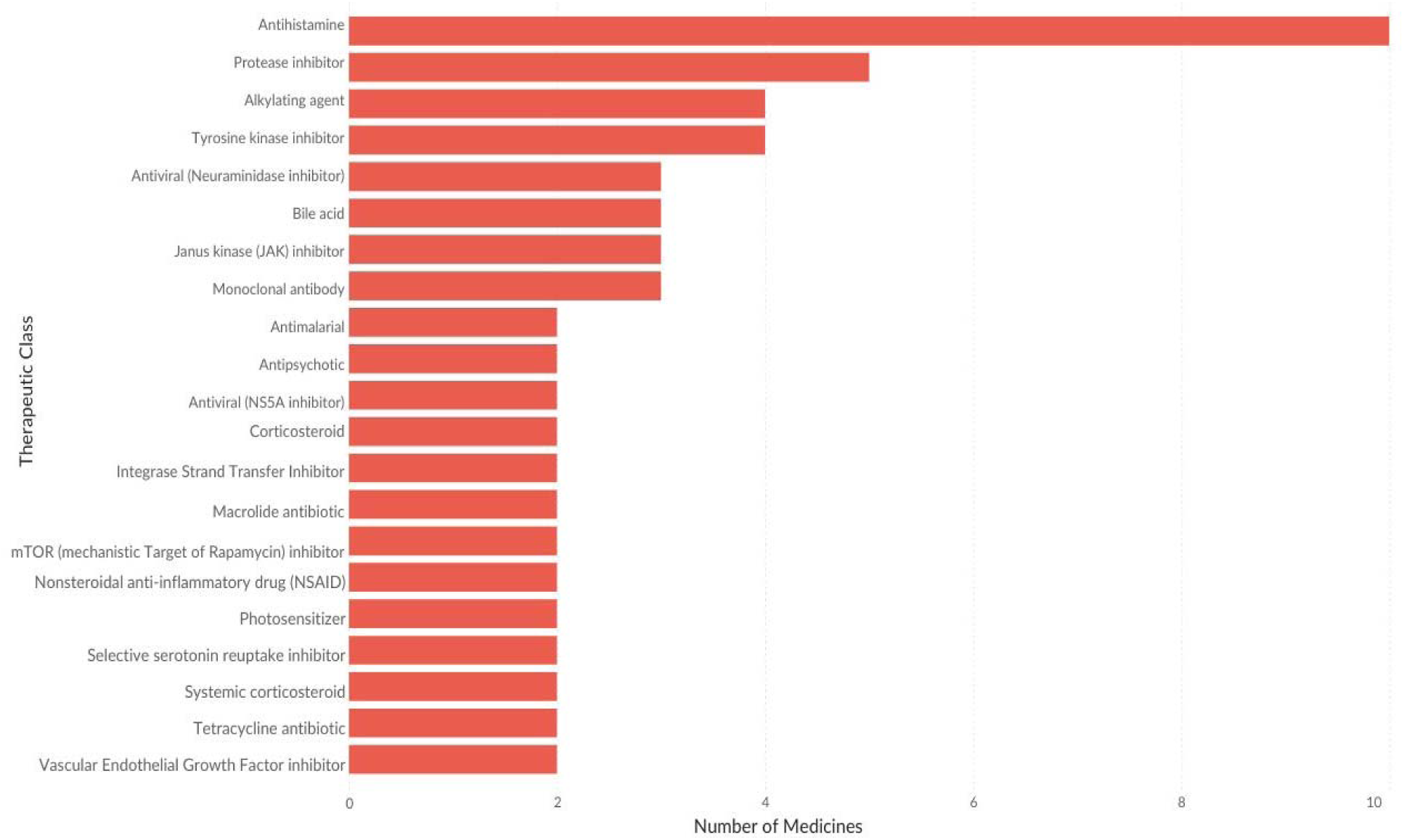
Number of identified repurposed technologies per therapeutic class for the treatment of MERS, SARS, and COVID-19. Only therapeutic classes with two or more technologies are shown.

### REPURPOSED TECHNOLOGIES IN ACTIVE CLINICAL DEVELOPMENT

The search conducted on ClinicalTrials.gov revealed that, among the viral diseases of interest, only COVID-19 and influenza are currently the focus of active interventional trials evaluating repurposed antiviral medicines.

Our analysis identified 30 unique interventional clinical trials, from which 58 technologies were retrieved (see Supplementary Tables 6 and 7). The discrepancy between the number of technologies and trials reflects the inclusion of multiple treatment arms within individual trials, encompassing both monotherapy and combination therapy approaches. In several instances, the investigated therapies involved combinations of medicines, where at least one was already approved for the target viral disease, and the other was repurposed from a non-related indication. These combinations represent novel therapeutic strategies aimed at enhancing efficacy or addressing resistance mechanisms.

Among the technologies identified, 18 were repurposed combination therapies, reflecting an interest in leveraging synergistic mechanisms of action to enhance antiviral efficacy or improve clinical outcomes. Only two of these combinations were being investigated for the treatment of influenza. The first combined oseltamivir, a neuraminidase inhibitor approved for influenza, with sirolimus, an mTOR (mammalian target of rapamycin) inhibitor typically used as an immunosuppressant; this strategy combines direct antiviral activity with modulation of host immune pathways. The second combination therapy paired oseltamivir with N-acetylcysteine, a mucolytic and antioxidant agent with potential to reduce oxidative stress and inflammation, thereby complementing antiviral activity. Both influenza-focused combination therapies were in phase III clinical development, indicating they are at an advanced stage of evaluation (Figure 7) (full details available in Supplementary Table 6).

**Figure 7.**
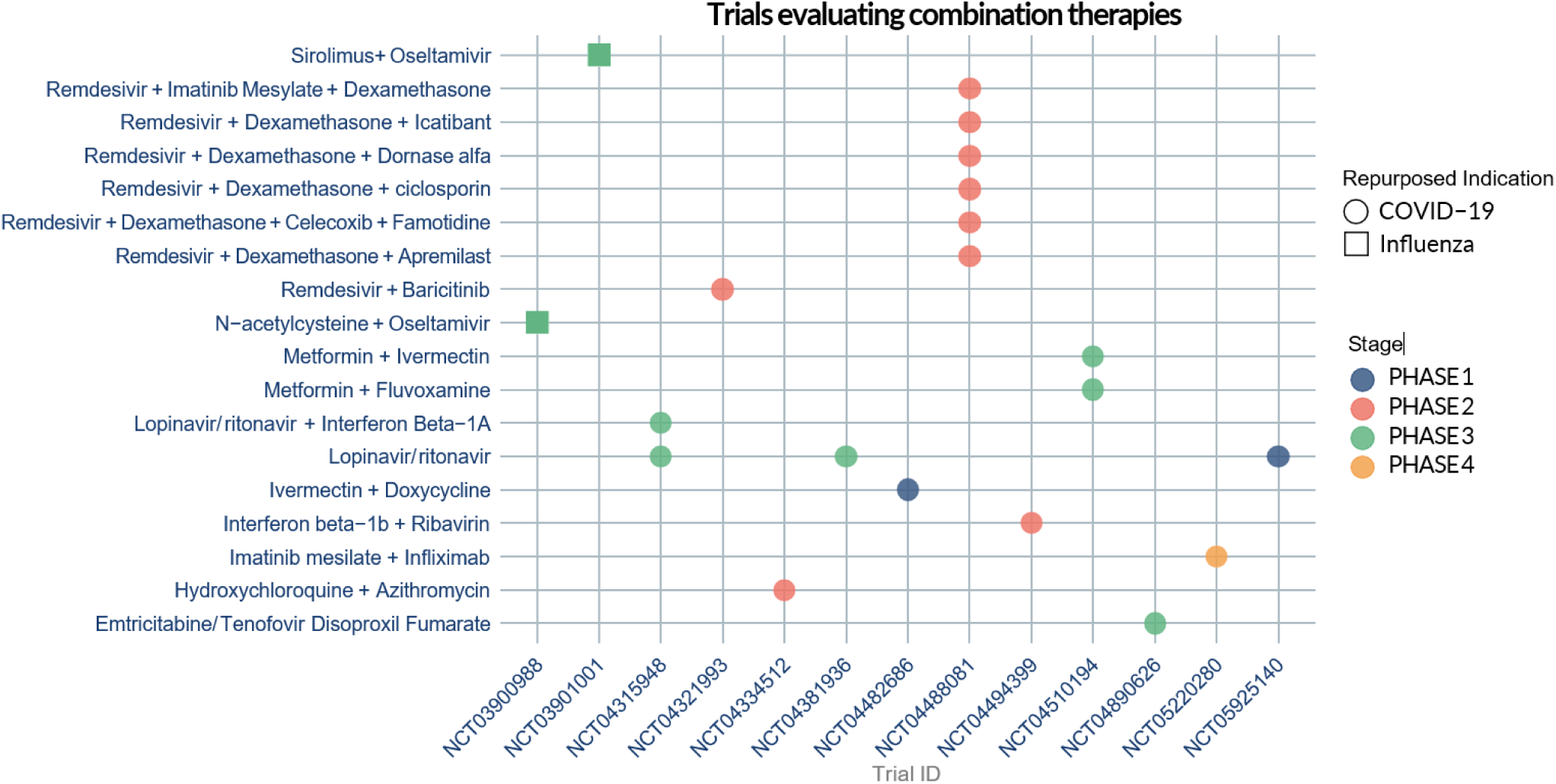
Graph showing the repurposed combination therapies in clinical development to treat COVID-19 and influenza

In contrast, combination therapies under investigation for COVID-19 were more diverse. The regimens included direct-acting antivirals (e.g. remdesivir), corticosteroids (e.g. dexamethasone), and immunomodulators such as Janus kinase inhibitors (e.g. baricitinib), among others. Of these COVID-19-focused combination therapies, six were in phase III clinical development, indicating substantial progress towards potential clinical adoption (Figure 7) (full details available in Supplementary Table 7). This diversity of treatment strategies underscores the complex pathophysiology of COVID-19 and highlights the importance of multidimensional approaches that can address both viral and host-mediated drivers of disease.

A total of forty repurposed monotherapies were identified across the included clinical trial records, with some, such as ivermectin and hydroxychloroquine, being evaluated in multiple studies. While a broad range of monotherapy candidates is under investigation for COVID-19, the pipeline for influenza appears far more limited. Only two repurposed medicines were identified as being in development for influenza: molnupiravir, a direct-acting antiviral currently in phase II clinical development, and dexamethasone sodium phosphate, a corticosteroid undergoing phase I evaluation (Figure 8) (full details available in Supplementary Table 6).

**Figure 8.**
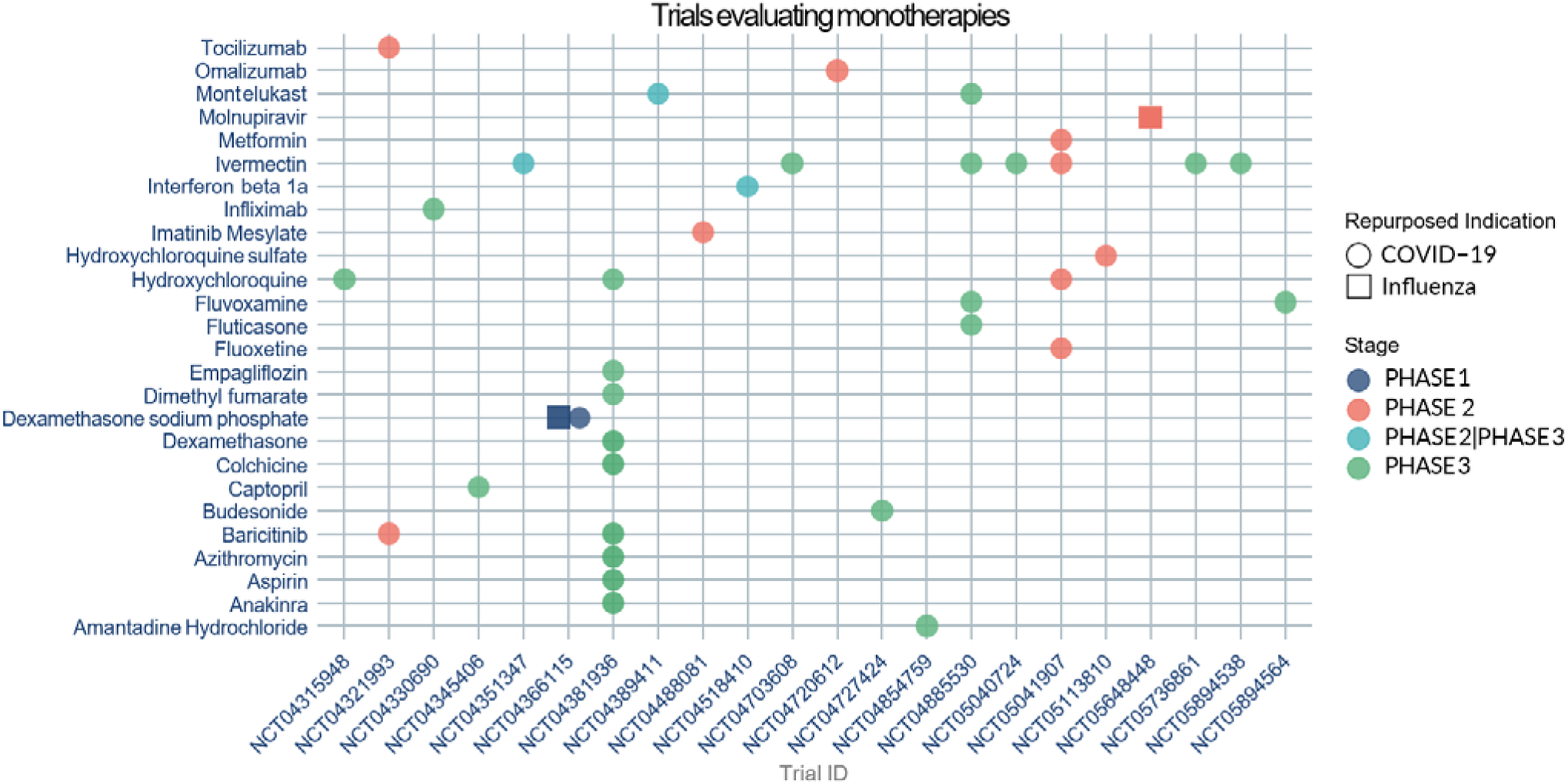
Graph showing the repurposed monotherapies in clinical development to treat COVID-19 and influenza

For COVID-19, the monotherapies encompassed a diverse range of pharmacological classes, reflecting strategies aimed at inhibiting viral replication and modulating host immune responses. These included interleukin-6 (IL-6) receptor inhibitors (e.g. tocilizumab), monoclonal antibodies (e.g. omalizumab), biguanides (e.g. metformin), and leukotriene receptor antagonists (e.g. montelukast), among others. Importantly, 23 of these monotherapies are in phase III clinical development, indicating substantial progress towards potential clinical adoption (Figure 8) (full details available in Supplementary Table 7). This diversity of mechanisms highlights the multifactorial nature of COVID-19 pathogenesis and the need for a broad therapeutic approach to address various stages of infection, mitigate complications, and improve patient outcomes.

## Discussion

This work provides a comprehensive horizon scan of repurposed antiviral medicines, either under investigation or proposed, for a defined set of viruses with pandemic potential. By synthesising evidence from published literature and registered clinical trials, we identified a diverse landscape of repurposing activity, which represent novel uses for medicines already licensed in the UK or EU. The identified technologies included medicines traditionally classified as antivirals as well as medicines from other therapeutic classes that possess known or potential antiviral activity. Some technologies involved combining medicines already licensed for a specific viral disease with others not yet approved for that indication, creating novel therapeutic strategies.

Clinical trial activity was observed to be concentrated exclusively on COVID-19 and influenza, with no active interventional trials identified for repurposed antiviral medicines targeting Ebola, Marburg, mpox, MERS, or SARS. The excluded trials for these viral diseases primarily reflected efforts to develop novel (unlicensed) therapeutics, a stronger emphasis on preventive interventions, or further evaluations of treatments already approved for these viral diseases. Although innovation through new drug development is essential, repurposing existing licensed medicines offers a pragmatic and potentially faster route to expand therapeutic options in the face of future pandemics.

The greatest concentration of repurposing activity was observed for COVID-19, reflecting the unprecedented global research mobilisation following the SARS-CoV-2 pandemic. A wide array of pharmacological classes was identified, targeting different aspects of COVID-19 pathogenesis, including direct-acting antivirals, host-modulatory agents such as interleukin-6 inhibitors, and immunomodulators. The range of mechanisms observed highlights the multifactorial nature of SARS-CoV-2 infection and the need for broad therapeutic strategies capable of addressing viral replication, inflammation, and downstream complications. A total of 29 repurposed technologies, encompassing both monotherapies and combination regimens, had progressed to phase III clinical evaluation, representing significant advancement towards potential clinical adoption and strengthening preparedness for future pandemic responses. In addition, 18 technologies were undergoing phase II evaluation, highlighting a robust mid-stage development pipeline with the potential to further expand therapeutic options. Therapeutic exploration for SARS and MERS was underrepresented; although historically significant, these viruses have received less recent research attention, likely reflecting a shift in focus towards SARS-CoV-2 as the more immediate coronavirus threat.

For influenza, a long-standing seasonal infection, the identified technologies targeted either direct viral inhibition or modulation of the host immune response. Only two repurposed monotherapies were in active clinical development for influenza: molnupiravir, a direct-acting antiviral in phase II trials, and dexamethasone, a corticosteroid undergoing phase I evaluation. In addition, two novel combination therapies involving oseltamivir (currently approved for influenza) were identified. Both combination therapies were in phase III clinical development, indicating substantial progress towards potential adoption into clinical practice.

For mpox, we identified 35 technologies from literature; two involved combination therapies involving tecovirimat (currently the only medicine approved for mpox) paired with other medicines licensed for other indications other than mpox. The exclusive approval of tecovirimat underscores the need for alternative options, especially in cases of resistance or treatment failure. The diversity of mechanisms among candidate therapies reflects growing interest in host-targeted approaches as complementary strategies. No clinical trials for mpox met the inclusion criteria, as they either focused on prophylactic vaccines, unlicensed medicines, or further evaluations of tecovirimat, which is already approved for mpox.

Therapeutic development for Ebola and Marburg viruses remains limited. For Ebola, 14 technologies were identified, while only two were reported for Marburg, all of which were extracted from literature. No clinical trials met the inclusion criteria for either virus, as existing studies primarily focused on prophylactic vaccines or involved unlicensed medicines.

A key feature of this scan was its focus on UK- or EU-licensed medicines with potential for repositioning. By excluding unlicensed medicines, the findings highlight opportunities for the strategic reuse of existing medicines and targeted support for ongoing repurposing efforts. The findings could inform future adaptive trial platforms and emergency use authorisations during pandemics.

Despite its strengths, this scan has limitations. The exclusion of non-English articles and reliance on a single literature database and clinical trials registry may have omitted relevant studies. The emphasis on recent publications (from 2022 onwards) helped capture timely innovations but may have missed earlier repurposing efforts still relevant to current development. Additionally, the analysis focused on medicines with existing UK or EU licences, which may not reflect the full global pipeline of repurposing candidates.

## Implications and Future Directions

The findings of this scan highlight the potential of existing licensed medicines for repurposing against emerging and re-emerging viral threats. Repurposing existing direct-acting antivirals or other pharmacological classes with potential antiviral activities offers a potentially rapid, cost-effective means of expanding treatment options in future outbreaks, provided that sufficient preclinical and clinical data support their use.

Indirect-acting antiviral medicines or host-targeted therapies, which modulate host pathways essential for viral replication or pathogenesis, represent a promising strategy to overcome challenges related to viral mutation and drug resistance. These medicines may offer broad-spectrum activity and sustained efficacy across different viral variants. Repurposing these medicines could significantly expand treatment options during the early stages of a viral outbreak, when virus-specific therapies are unavailable or still in development.

Looking ahead, collaboration among researchers, research funders, pharmaceutical industry, policymakers, public health agencies, and regulatory bodies will be essential to improve pandemic preparedness. Such coordination is also vital for accelerating the regulatory evaluation and adoption of repurposed medicines, ensuring that safe and effective therapeutic options can be rapidly mobilised in future public health emergencies.

## Author contributions

SA drafted the manuscript. AI conducted the literature and clinical trial searches. SA, RP, JN, and KB carried out data screening and extraction. SA and RP analysed the data. GN reviewed and provided feedback on the final draft of the manuscript. All authors provided critical feedback and helped refine the manuscript.

## Acknowledgment

We sincerely thank Katie Twentyman (NIHR Innovation Observatory) for her contribution to this work, particularly in creating some of the visuals included in this manuscript.

## Data Availability Statement

All data supporting the findings of this study are provided in the supplementary material.

## Appendix A

### Literature search

#### Search conducted in Embase bibliographic database on 8^th^ April 2025

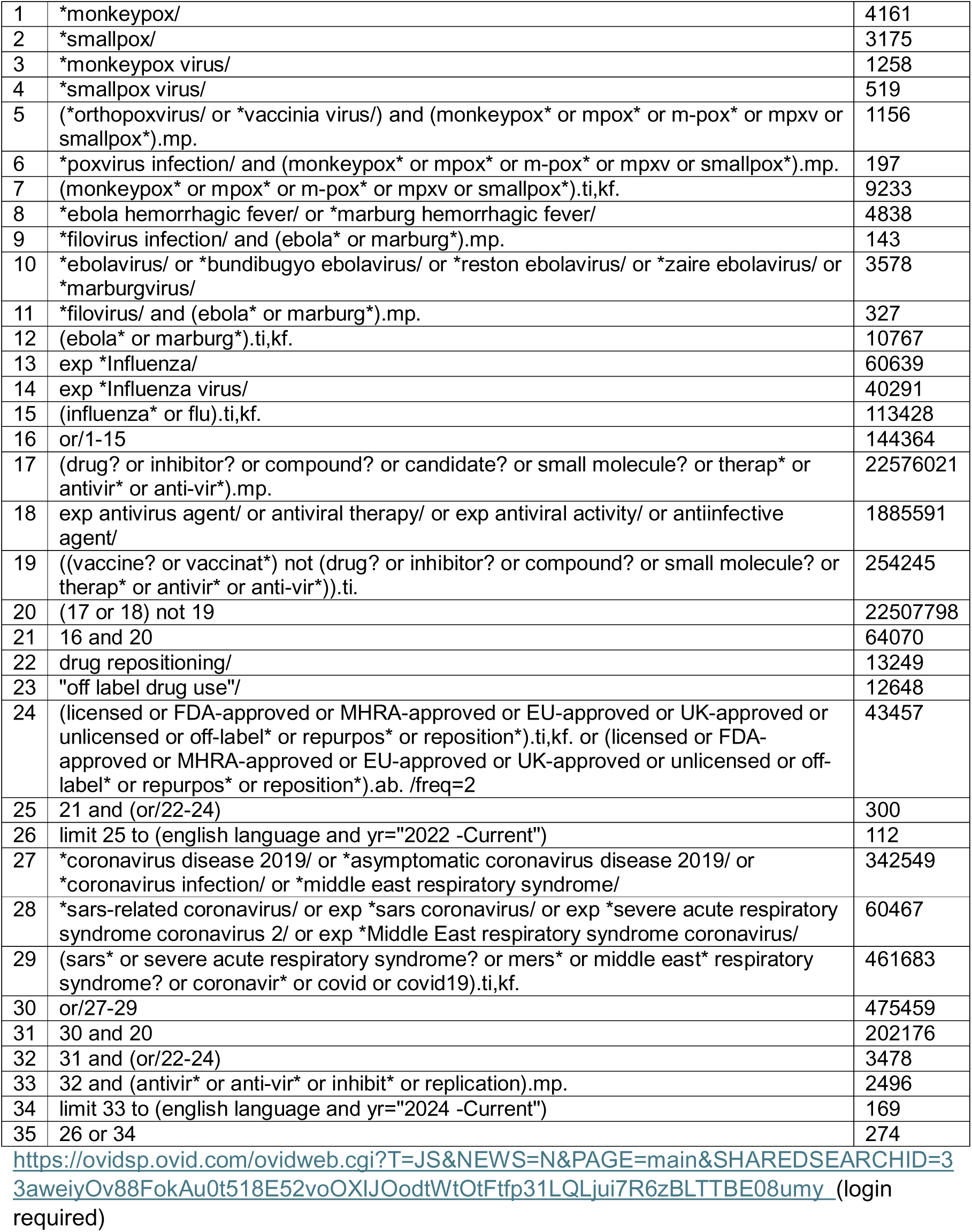

