## Supplementary Material for "Repurposed antiviral medicines for potential pandemic viruses: A horizon scan"

**Supplementary Tables Detailing the Identified Repurposed Technologies (Tables 1- 7)**

**Supplementary Table 1 –** **Candidates for the treatment of Ebola**

| **Medicines** | **Therapeutic Class (A-Z)** | **UK/EU Approved Indication** | **Cancer Y/N** | **References** |
| --- | --- | --- | --- | --- |
| Trifluoperazine | Antipsychotic | Depressive symptoms secondary to anxiety, Agitation, Nausea and vomiting, Schizophrenia and in other psychoses, Severe psychomotor agitation and of dangerously impulsive behaviour | No | Almeida-Pinto 2024,^1^ Zeidan 2024^2^ |
| Prochlorperazine | Antipsychotic and antiemetic | Vertigo due to Ménière's disease, Labyrinthitis and other causes, Nausea and vomiting, Migraine | No | Almeida-Pinto 2024,^1^ Zeidan 2024^2^ |
| Glecaprevir | Antiviral (NS3/4A protease inhibitor) | Chronic hepatitis C | No | Broni 2023^3^ |
| Ledipasvir | Antiviral (NS5A inhibitor) | Chronic hepatitis C | No | Broni 2023^3^ |
| Velpatasvir | Antiviral (NS5A inhibitor) | Chronic hepatitis C | No | Broni 2023^3^ |
| Remdesivir | Antiviral (RNA polymerase inhibitor) | Covid-19 | No | Barghash 2024,^4^ Nascimento 2022^5^ |
| Teicoplanin | Glycopeptide antibiotic | Complicated skin and soft tissue infections, Bone and joint infections, Hospital acquired pneumonia, Community acquired pneumonia, Complicated urinary tract infections, Infective endocarditis, Peritonitis associated with continuous ambulatory peritoneal dialysis, Clostridium difficile infection | No | Barghash 2024,^4^ Espano 2024^6^ |
| Simvastatin | HMG-CoA reductase inhibitor | Hypercholesterolaemia, Cardiovascular prevention | No | Almedia-Pinto 2024^1^ |
| Raloxifene | Selective oestrogen receptor modulator | Osteoporosis | No | Almeida-Pinto 2024,^1^ Nascimento 2022^5^ |
| Tamoxifen | Selective oestrogen receptor modulator | Anovulatory infertility, Breast cancer | Yes | Almeida-Pinto 2024,^1^ Nascimento 2022^5^ |
| Toremifene | Selective oestrogen receptor modulator | Breast cancer | Yes | Almeida-Pinto 2024,^1^ Nascimento 2022^5^ |
| Fluoxetine | Selective serotonin reuptake inhibitor | Bulimia nervosa, Major depressive disorders, obsessive-compulsive disorder | No | Kummer 2022^7^ |
| Eltrombopag | Thrombopoietin receptor agonist | Acquired aplastic anaemia, Hepatitis C, Primary immune thrombocytopenia | No | Broni 2023^3^ |
| Itraconazole | Triazole antifungal | Aspergillosis, Candidosis, Cryptococcosis, Histoplasmosis | No | Almeida-Pinto 2024,^1^ Kummer 2022,^7^ Vanmechelen 2022^8^ |

**Supplementary Table 2 – Candidates for the treatment of Marburg**

| **Medicines** | **Therapeutic Class** | **UK/EU Approved Indications** | **Cancer Y/N** | **References** |
| --- | --- | --- | --- | --- |
| Remdesivir | Antiviral (RNA polymerase inhibitor) | COVID-19 | No | Martins 2025^9^ |
| Bictegravir | Integrase Strand Transfer Inhibitor | Human immunodeficiency virus | No | Singh 2025^10^ |

**Supplementary Table 3 – Candidates for the treatment of Influenza**

| **Medicines** | **Therapeutic Class (A-Z)** | **UK/EU Approved Indications** | **Cancer Y/N** | **References** |
| --- | --- | --- | --- | --- |
| Telmisartan | Angiotensin II receptor blocker | Hypertension, Cardiovascular prevention in manifest atherothrombotic cardiovascular disease or patients with type 2 diabetes mellitus with documented target organ damage | No | Taye 2023^11^ |
| Enalapril | Angiotensin-Converting Enzyme inhibitor | Heart failure, Hypertension, Cardiovascular event prevention | No | Ghimire 2022^12^ |
| Ranolazine | Antianginal | Add on treatment for stable angina pectoris | No | Taye 2024^11^ |
| Idarubicin | Antineoplastic | Acute non-lymphocytic leukaemia, Advanced breast cancer, Second line relapsed acute lymphoblastic leukaemia | Yes | Bordoloi 2023^13^ |
| Lurasidone | Antipsychotic | Schizophrenia | No | Mtambo 2022^14^ |
| Dextromethorphan | Antitussive | Cough | No | Xie 2022^15^ |
| Molnupiravir | Antiviral (RNA polymerase inhibitor) | COVID-19 | No | Padey 2024^16^ |
| Dabrafenib | BRAF kinase inhibitor | Melanoma | Yes | Meineke 2022^17^ |
| Diltiazem | Calcium channel blocker | Angina pectoris, Hypertension | No | Padey 2024^16^ |
| Ofloxacin | Fluoroquinolone antibiotic | Acute pyelonephritis and complicated urinary tract infections, non-gonococcal urethritis and cervicitis, Gonococcal urethritis and cervicitis due to susceptible *Neisseria gonorrhoeae*, Acute exacerbations of chronic obstructive pulmonary disease, Uncomplicated cystitis, Urethritis | No | Taye 2023^11^ |
| Maraviroc | HIV entry inhibitor | In combination with other medicinal products for human immunodeficiency virus | No | Xie 2022^15^ |
| Ivabradine | Hyperpolarisation-activated cyclic nucleotide-gated (HCN) channel blockers. | Chronic stable angina pectoris, Chronic heart failure | No | Ghimire 2022^12^ |
| Miconazole | Imidazole antifungal | Mycotic infections of the skin and superinfections due to Gram positive bacteria | No | Bordoloi 2023^13^ |
| Econazole | Imidazole antifungal | Candida/yeast infections, Vulvovaginitis and mycotic balanitis, Dermatophytosis and pityriasis versicolor | No | Bordoloi 2023^13^ |
| Tioconazole | Imidazole antifungal | Nail infections | No | Bordoloi 2023^13^ |
| Everolimus | mTOR (mechanistic Target of Rapamycin) inhibitor | Neuroendocrine tumours of pancreatic origin, Neuroendocrine tumours of gastrointestinal or lung origin, Renal cell carcinoma | Yes | Sun 2025^18^ |
| Regorafenib | Multikinase inhibitor | Colorectal cancer, Gastrointestinal stromal tumours, Hepatocellular carcinoma | Yes | Meineke 2022^17^ |
| Ribavirin | Nucleoside antiviral (RNA synthesis inhibitor) | In combination with other drugs: Chronic hepatitis c | No | Li 2023^19^ |
| Metoclopramide | Prokinetic and antiemetic | Nausea and vomiting | No | Taye 2023^11^ |
| Eltrombopag | Thrombopoietin receptor agonist | Primary immune thrombocytopenia, Hepatitis c, Acquired aplastic anaemia | No | Mtambo 2022^14^ |
| Larotrectinib | Tropomyosin receptor kinase inhibitor | Solid tumours | Yes | Meineke 2022^17^ |
| Afatinib | Tyrosine kinase inhibitor | Non-small cell lung cancer | Yes | Meineke 2022^17^ |
| Avapritinib | Tyrosine kinase inhibitor | Gastrointestinal stromal tumour, Systemic mastocytosis | Yes | Meineke 2022^17^ |
| Neratinib | Tyrosine kinase inhibitor | Breast cancer | Yes | Meineke 2022^17^ |
| Tucatinib | Tyrosine kinase inhibitor | Breast cancer | Yes | Mtambo 2022^14^ |
| Bosutinib | Tyrosine kinase inhibitor | Chronic myeloid leukaemia | Yes | Xie 2022^15^ |

**Supplementary Table 4 – Candidates for the treatment of Monkeypox (mpox)**

| **Medicines** | **Therapeutic Class (A-Z)** | **UK/EU Approved Indications** | **Cancer Y/N** | **References** |
| --- | --- | --- | --- | --- |
| Chloramphenicol | Amphenicol antibiotic | Bacterial conjunctivitis, Typhoid, Meningitis, Gram-negative and Gram-positive organisms | No | Podduturi 2024^20^ |
| Mitoxantrone | Anthracenedione antineoplastic | Breast cancer, non-Hodgkin’s lymphoma, Acute myeloid leukaemia | No | Preet 2022^21^ |
| Doxorubicin | Anthracycline antineoplastic | Breast cancer, Sarcoma, Small-cell carcinoma of the lung, Hodgkin disease or non-Hodgkin lymphoma | Yes | Sahoo 2023^22^ |
| Fenofibrate + tecovirimat | Antilipemic agent + Antiviral | Hypertriglyceridaemia, Mixed hyperlipidaemia \| Smallpox, Monkeypox, Cowpox | No | Vuorio 2022^23^ |
| Chloroquine diphosphate | Antimalarial | Active rheumatoid arthritis, Malaria, Amoebic hepatitis and abscess, Discoid and systemic lupus erythematosus | No | Horton 2025^24^ |
| Baloxavir | Antiviral (Cap-dependent endonuclease inhibitor) | Influenza | No | Hashemi 2024,^25^ Horton 2025,^24^ Rabaan 2024^26^ |
| Glecaprevir | Antiviral (NS3/4A protease inhibitor) | Chronic hepatitis C | No | Li 2023^27^ |
| Cidofovir | Antiviral (Nucleotide analogue DNA polymerase inhibitor) | Cytomegalovirus retinitis | No | Aldhaeefi 2023,^28^ Bhattacharjee 2024,^29^ Bojkova 2023,^30^ Borkotoky 2024,^31^ Ezat 2023,^32^ Horton 2025,^24^ Islam 2022,^33^ Lam 2022,^34^ Rejinold 2025,^35^ Shannon 2025,^36^ Vuorio 2022,^23^ Yousaf 2025^37^ |
| Tecovirimat + mycophenolate | Antiviral + Immunosuppressant | Smallpox, Monkeypox and Cowpox \| Prophylaxis of acute transplant rejection | No | Borkotoky 2024,^31^ Witwit 2025^38^ |
| Ibandronate | Bisphosphonate | Osteoporosis | No | Podduturi 2024^20^ |
| Lumacaftor | Cystic fibrosis transmembrane conductance regulator corrector | Cystic fibrosis | No | Dutt 2023,^39^ Khan 2024,^40^ Li 2023^27^ |
| Eravacycline | Fluorocycline antibiotic | Complicated intra-abdominal infections | No | Alandijany 2023^41^ |
| Bictegravir | Integrase Strand Transfer Inhibitor | In combination with other drugs for the treatment of Human immunodeficiency virus | No | Li 2023^27^ |
| cabotegravir | Integrase Strand Transfer Inhibitor | Prophylaxis Human immunodeficiency virus | No | Li 2023,^27^ Patel 2023^42^ |
| Dolutegravir | Integrase Strand Transfer Inhibitor | In combination with other drugs for the treatment of Human immunodeficiency virus | No | Li 2023,^27^ Lythgoe 2024,^43^ Sahoo 2023^22^ |
| Elvitegravir | Integrase Strand Transfer Inhibitor | Human immunodeficiency virus | No | Patel 2023^42^ |
| Baricitinib | Janus kinase (JAK) inhibitor | Rheumatoid arthritis, Atopic dermatitis, Alopecia areata, Juvenile idiopathic arthritis | No | Rabaan 2024^26^ |
| Capmatinib | MET tyrosine kinase inhibitor | Non-small cell lung cancer | Yes | Li 2023,^27^ Srivastava 2024^44^ |
| Tepotinib | MET tyrosine kinase inhibitor | Non-small cell lung cancer | No | Abduljalil 2023,^41^ Li 2023^27^ |
| Tipranavir | Non-peptidic protease inhibitor | Human immunodeficiency virus | No | Lythgoe 2024,^43^ Patel 2023,^42^ Sahoo 2023^22^ |
| Ribavirin | Nucleoside antiviral (RNA synthesis inhibitor) | In combination with other drugs for chronic hepatitis C | Yes | Ezat 2023,^32^ Hashemi 2024^25^ |
| Zidovudine | Nucleoside reverse transcriptase inhibitor | Human immunodeficiency virus | Yes | Bhattacharjee 2024,^29^ Patel 2023,^42^ Rabaan 2024^26^ |
| Naldemedine | Opioid antagonist | Opioid-induced constipation | Yes | Srivastava 2024^44^ |
| Cannabidiol | Phytocannabinoid | Seizures associated with tuberous sclerosis complex, Lennox-Gastaut syndrome, Multiple sclerosis | No | Paul 2024^45^ |
| Amphotericin | Polyene antifungal | Mycoses, Visceral leishmaniasis, Fungal infections, Candidiasis | No | Peruzzu 2023^46^ |
| Mefloquine hydrochloride | Quinoline antimalarial | P. falciparum malaria | No | Horton 2025^24^ |
| Cefiderocol | Siderophore cephalosporin antibiotic | Gram-negative infections | No | Sahoo 2023^22^ |
| Fostamatinib | Spleen tyrosine kinase inhibitor | Chronic immune thrombocytopenia | No | Paul 2024,^45^ Saha 2024^47^ |
| Doxycycline | Tetracycline antibiotic | Respiratory tract infections, Urinary tract infections, Sexually transmitted infections, Skin infections | No | Yousaf 2025^37^ |
| Adalimumab | Tumour necrosis factor (TNF) inhibitor | Ankylosing spondylitis, Psoriatic arthritis, Hidradenitis suppurativa, Crohn's disease | No | Rabaan 2024^26^ |
| Etanercept | Tumour necrosis factor (TNF) inhibitor | Psoriatic arthritis, Axial spondyloarthritis, Plaque psoriasis | No | Rabaan 2024^26^ |
| Infliximab | Tumour necrosis factor (TNF) inhibitor | Crohn's disease, Ulcerative colitis, Ankylosing spondylitis, Psoriatic arthritis, Psoriasis | Yes | Rabaan 2024^26^ |
| Imatinib | Tyrosine kinase inhibitor | Chronic myelogenous leukemia, Acute lymphoblastic leukemia, Myelodysplastic/myeloproliferative diseases | No | Dutt 2023,^39^ Ezat 2023,^32^ Rabaan 2024^26^ |
| Nilotinib | Tyrosine kinase inhibitor | Chronic myelogenous leukemia | No | Khan 2024,^40^ Li 2023^27^ |
| Ponatinib | Tyrosine kinase inhibitor | Chronic myeloid leukaemia, Acute lymphoblastic leukaemia | Yes | Khan 2024^40^ |

**Supplementary Table 5 – Candidates for the treatment of MERS, SARS, COVID-19**

| **Repurposed indication** | **Medicines** | **Therapeutic Class (A-Z)** | **UK/EU Approved Indications** | **Cancer Y/N** | **References** |
| --- | --- | --- | --- | --- | --- |
| COVID | Pyridostigmine bromide | Acetylcholinesterase inhibitor | Myasthenia gravis, Paralytic ileus, post-operative urinary retention | No | Jaimes-Castelan 2024^48^ |
| COVID | Disulfiram | Aldehyde dehydrogenase inhibitor | Alcohol abuse | No | Boulon 2024,^49^ Mia 2024,^50^ Nazir 2024^51^ |
| COVID | Busulfan | Alkylating agent | Chronic myeloid leukaemia, Polycythaemia vera, Thrombocythemia, Myelofibrosis | Yes | Alzahrani 2024^52^ |
| COVID | Carboplatin | Alkylating agent | Ovarian carcinoma of epithelial origin, small cell lung carcinoma | Yes | Alzahrani 2024^52^ |
| COVID | Cisplatin | Alkylating agent | Testicular cancer, Bladder cancer, Ovarian cancer, Head and neck cancer, non-small cell lung cancer, small cell lung cancer | Yes | Brun 2025^53^ |
| COVID | Ifosfamide | Alkylating agent | Malignant disease | Yes | Alzahrani 2024^52^ |
| COVID | Dexmedetomidine | Alpha-2 receptor agonist | Sedation in non-intubated patients | No | Kumawat 2024^54^ |
| COVID | Telmisartan | Angiotensin II receptor blocker | Hypertension, Cardiovascular prevention | No | Aviles-Alia 2024^55^ |
| COVID | Fosinopril | Angiotensin-Converting Enzyme inhibitor | Hypertension, Heart failure | No | Metwaly 2024^56^ |
| COVID | Glycopyrronium + formoterol fumarate | Anticholinergic (long-acting muscarinic antagonist) + long-acting beta-2 agonist | Chronic obstructive pulmonary disease | No | Enyeji 2024^57^ |
| COVID | Enoxaparin | Anticoagulant | Venous thromboembolic disease, Deep vein thrombosis, Prevention of thrombus formation, Myocardial infarction | No | Jaimes-Castelan 2024^48^ |
| COVID | Valproic acid | Antiepileptic | Epilepsy | No | Alzahrani 2024^52^ |
| COVID | Acrivastine | Antihistamine | Rhinitis, Urticaria | No | Zhong 2024^58^ |
| COVID | Azelastine | Antihistamine | Seasonal allergic conjunctivitis, Seasonal allergic rhinitis | No | Zhong 2024^58^ |
| COVID | Bilastine | Antihistamine | Allergic rhino-conjunctivitis, Urticaria. | No | Hamdan 2024^59^ |
| COVID | Desloratadine | Antihistamine | Allergic rhinitis, Urticaria | No | Zhong 2024^58^ |
| COVID | Diphenhydramine | Antihistamine | Allergic conditions of the skin | No | Zhong 2024^58^ |
| COVID | Fexofenadine | Antihistamine | Allergic rhinitis, Urticaria | No | Hamdan 2024^59^ |
| COVID | Loratadine | Antihistamine | Allergic rhinitis, Urticaria | No | Zhong 2024^58^ |
| COVID | Promethazine | Antihistamine | Allergic rhinitis, Urticaria, Anaphylactic reactions to drugs/ foreign proteins, Antiemetic, Insomnia and paediatric sedation, | No | Zeidan 2024^2^ |
| COVID | Rupatadine | Antihistamine | Allergic rhinitis, Urticaria | No | Hamdan 2024^59^ |
| COVID | Triprolidine | Antihistamine | In combination with other drugs for: Upper respiratory tract disorders | No | Zhong 2024^58^ |
| COVID | Chloroquine diphosphate | Antimalarial | Active rheumatoid arthritis, Malaria, Amoebic hepatitis and abscess, Discoid and systemic lupus erythematosus | No | Chavda 2024,^60^ Enyeji 2024,^57^ Islam 2024,^61^ Kumawat 2024,^54^ Mia 2024^50^ |
| COVID | Pyronaridine + Artesunate | Antimalarial | Malaria | No | Abla 2024^62^ |
| COVID | Hydroxychloroquine | Antimalarial and immunomodulatory agent | Rheumatoid arthritis, Discoid and systemic lupus erythematosus, Dermatological conditions, Lupus erythematosus | No | Chavda 2024,^60^ Enyeji 2024,^57^ Jaimes-Castelan 2024,^48^ Kumawat 2024,^54^ Martins 2024,^63^ Mia 2024,^50^ Singh 2024,^64^ Islam 2024^61^ |
| COVID | Rifampicin | Antimicrobial | Tuberculosis, leprosy, Brucellosis, Legionnaires Disease, Staphylococcal infections, Meningococcal meningitis, Haemophilus influenzae | No | Chakraborty 2024^65^ |
| COVID, MERS, SARS | Ivermectin | Antiparasitic | Anguillulosis, Microfilaraemia, Scabies, Inflammatory lesions of rosacea | No | Chavda 2024,^60^ Enyeji 2024,^57^ Gossen 2024,^66^ Jaimes-Castelan 2024,^48^ Mawazi 2024,^67^ Mia 2024,^50^ Pereira 2024,^68^ Singh 2024,^64^ Xu 2024^69^ |
| COVID | Pyrimethamine | Antiprotozoal | In combination with a synergistic agent for: Toxoplasmosis | No | Godde 2024^70^ |
| COVID, MERS, SARS | Chlorpromazine | Antipsychotic | Schizophrenia and other psychoses, Mania and hypomania, Anxiety, Psychomotor agitation, Excitement, Violent or dangerously impulsive behaviour, Intractable hiccup, Nausea and vomiting in terminal illness, Induction of hypothermia, Childhood schizophrenia, Autism | No | Zeidan 2024,^2^ Barghash 2024^4^ |
| COVID | Haloperidol | Antipsychotic | Schizophrenia and schizoaffective disorder, Delirium, Bipolar, Acute psychomotor agitation, Persistent aggression and psychotic symptoms, Tourette's syndrome | No | Boulon 2024,^49^ Zeidan 2024^2^ |
| COVID, MERS, SARS | Oseltamivir | Antiviral (Neuraminidase inhibitor) | Influenza | No | Gao 2024,^71^ Jaimes-Castelan 2024,^48^ Marinho 2025,^72^ Mia 2024,^50^ Waseem 2024,^73^ Barghash 2025^4^ |
| COVID, MERS, SARS | Zanamivir | Antiviral (Neuraminidase inhibitor) | Influenza A and B | No | Gao 2024,^71^ Mia 2024,^50^ Waseem 2024,^73^ Barghash 2024^4^ |
| COVID | Zanamivir | Antiviral (Neuraminidase inhibitor) | Influenza A and B | No | Gao 2024,^71^ Mia 2024,^50^ Waseem 2024,(71) Barghash 2024^4^ |
| COVID | Elbasvir | Antiviral (NS5A inhibitor) | In combination with other drugs for: Chronic hepatitis C | No | Ahmad 2024^74^ |
| COVID | Ledipasvir | Antiviral (NS5A inhibitor) | Chronic hepatitis C | No | Gao 2024,^71^ Pereira 2024^68^ |
| COVID | Sofosbuvir | Antiviral (NS5B polymerase inhibitor) | In combination with other drugs for: Chronic hepatitis C | No | Gao 2024,^71^ Kumawat 2024,^54^ Metwaly 2024,^56^ Singh 2024^64^ |
| COVID, MERS, SARS | Amantadine | Antiviral and antiparkinsonian | Parkinson's disease | No | Barghash 2024^4^ |
| COVID | Zepatier (Elbasvir/Grazoprevir) | Antivirals (NS5A inhibitor + NS3/4A protease inhibitor) | Fixed dose combination for: Chronic hepatitis C | No | Ahmad 2024^74^, Pereira 2024^68^ |
| COVID | Epclusa | Antivirals (NS5B polymerase inhibitor + NS5A inhibitor) | Licensed combination= sofosbuvir/velpatasvir for: chronic hepatitis C | No | Boulon 2024^49^ |
| COVID | Venetoclax | BCL-2 (B-cell lymphoma 2) inhibitor | Chronic lymphocytic leukaemia | Yes | Xu 2024^69^ |
| COVID, MERS, SARS | Metformin | Biguanide | Type 2 Diabetes, Polycystic ovary syndrome | No | Singh 2024,^64^ Yip 2025,^75^ Barghash 2024^4^ |
| COVID | Chenodeoxycholic acid | Bile acid | Cerebrotendinous xanthomatosis | No | Fiorucci 2024^76^ |
| COVID | Ursodeoxycholic acid | Bile acid | Cholesterol stones, Hepatobiliary disorder associated with cystic fibrosis | No | Fiorucci 2024,^76^ Lee 2024^77^ |
| COVID | Vemurafenib | BRAF kinase inhibitor | Melanoma | Yes | Alzahrani 2024,^52^ Bogacheva 2024^78^ |
| COVID | Diltiazem | Calcium channel blocker | Angina pectoris, Hypertension | No | Padey 2024^16^ |
| COVID | Digoxin | Cardiac glycoside | Cardiac failure, Supraventricular arrhythmias | No | Alzahrani 2024,^52^ Boulon 2024,^49^ Pereira 2024^68^ |
| COVID | Budesonide | Corticosteroid | Seasonal allergic rhinitis | No | Enyeji 2024,^57^ Martins 2024^63^ |
| COVID | Hydrocortisone | Corticosteroid | Adrenal insufficiency, Haemorrhoids, Pruritus ani, Eczema, Allergic contact dermatitis, Irritant contact dermatitis, Stings/bug bites, Photodermatitis, Otitis externa, Intertrigo, Prurigo nodularis, Seborrhoeic dermatitis, Congenital adrenal hyperplasia, Severe bronchial asthma, Drug hypersensitivity reactions | No | Alzahrani 2024^52^ |
| COVID | Aliskiren | Direct renin inhibitor | Essential hypertension | No | Metwaly 2024^56^ |
| COVID | Estradiol | Estrogen hormone | Hormone replacement therapy, Prevention of osteoporosis | No | Aviles-Alia 2024,^55^ Xu 2024^69^ |
| COVID | Ionafarnib | Farnesyltransferase Inhibitor | Hutchinson-Gilford progeroid syndrome, Processing-deficient progeroid laminopathies | No | Khan 2025^79^ |
| COVID | Ofloxacin | Fluoroquinolone antibiotic | Acute pyelonephritis and complicated urinary tract infections, non-gonococcal urethritis and cervicitis, Gonococcal urethritis and cervicitis due to susceptible Neisseria gonorrhoea’s, Acute exacerbations of chronic obstructive pulmonary disease, Uncomplicated cystitis, Urethritis | No | Chakraborty 2024^65^ |
| COVID | Miglustat | Glucosylceramide synthase inhibitor | Gaucher disease, Niemann-Pick type C disease, Pompe disease | No | Brun 2025^53^ |
| COVID | Riluzole | Glutamate release inhibitor | Amyotrophic lateral sclerosis | No | Marquez-Monino 2025^80^ |
| COVID | Teicoplanin | Glycopeptide antibiotic | Complicated skin and soft tissue infections, Bone and joint infections, Pneumonia, Urinary tract infections, Infective endocarditis, Peritonitis, Clostridium difficile infection | No | Espano 2024,^6^ Mia 2024,^50^ Barghash 2024^4^ |
| COVID | Simvastatin | HMG-CoA reductase inhibitor | Hypercholesterolaemia, Mixed dyslipidaemia, Cardiovascular prevention | No | Alzahrani 2024^52^ |
| COVID | Imiquimod | Immune response modifier | External genital and perianal warts, Superficial basal cell carcinomas, Actinic keratoses on face or scalp | No | Voloudakis 2025,^81^ Waseem 2024^73^ |
| COVID | Azathioprine | Immunosuppressant | Inflammatory bowel disease, Severe active rheumatoid arthritis, Systemic lupus erythematosus, Dermatomyositis, polymyositis, Auto-immune chronic active hepatitis, Pemphigus vulgaris, Polyarteritis nodosa, Auto-immune haemolytic anaemia, Chronic refractory idiopathic thrombocytopenic purpura | No | Alzahrani 2024,^52^ Voloudakis 2025^81^ |
| COVID | Ciclesonide | Inhaled corticosteroid | Asthma | No | Enyeji 2024^57^ |
| COVID | Dolutegravir | Integrase Strand Transfer Inhibitor | In combination with other drugs for: Human Immunodeficiency Virus | No | Kasgari 2025^82^ |
| COVID | Raltegravir | Integrase Strand Transfer Inhibitor | In combination with other drugs for: Human Immunodeficiency Virus | No | Gao 2024,^71^ Mohamed 2025^83^ |
| COVID | Anakinra | Interleukin-1 receptor antagonist | Rheumatoid arthritis, Periodic fever syndromes, Still's disease | No | Chavda 2024,^60^ Jaimes-Castelan 2024,^48^ Mawazi 2024^67^ |
| COVID | Baricitinib | Janus kinase (JAK) inhibitor | Rheumatoid arthritis, Atopic dermatitis, Alopecia areata, Juvenile idiopathic arthritis | No | Chavda 2024,^60^ Jaimes-Castelan 2024,^48^ Low 2025,^84^ Rahmani 2024^85^ |
| COVID | Ruxolitinib | Janus kinase (JAK) inhibitor | Myelofibrosis, Polycythaemia vera, Graft versus host disease | No | Chavda 2024^60^ |
| COVID | Tofacitinib | Janus kinase (JAK) inhibitor | Active polyarticular juvenile idiopathic arthritis, Juvenile psoriatic arthritis | No | Jaimes-Castelan 2024^48^ |
| COVID | Montelukast | Leukotriene receptor antagonist | Asthma, Allergic rhinitis associated with asthma | No | Hamdan 2024^59^ |
| COVID | Azithromycin | Macrolide antibiotic | Bacterial sinusitis, Bacterial otitis media, Pharyngitis, Tonsillitis, Chronic bronchitis, Community acquired pneumonia, Skin and soft tissue infections, uncomplicated Chlamydia trachomatis urethritis and cervicitis, conjunctivitis (eye drops), pelvic inflammatory disease, sinusitis | No | Chavda 2024,^60^ Enyeji 2024,^57^ Mia 2024,^50^ Paroczai 2024,^86^ Singh 2024^64^ |
| COVID | Fidaxomicin | Macrolide antibiotic | Clostridioides difficile infections | No | Protic 2024^87^ |
| COVID | Trametinib | MEK (mitogen-activated extracellular signal-regulated kinase) inhibitor | In combination with other drugs for: Melanoma, Non-small cell lung cancer | Yes | Alzahrani 2024^52^ |
| COVID | Sarilumab | Monoclonal antibody | Rheumatoid arthritis, Polymyalgia rheumatica | No | Chavda 2024^60^ |
| COVID | Siltuximab | Monoclonal antibody | Multicentric Castleman's disease | No | Chavda 2024^60^ |
| COVID | Tocilizumab | Monoclonal antibody | Rheumatoid arthritis, Active systemic juvenile idiopathic arthritis, Giant cell arteritis | No | Chavda 2024,^60^ Jaimes-Castelan 2024,^48^ Kumawat 2024,^54^ Okeowo 2024,^88^ Papp 2024,^89^ Low 2025,^84^ Rahmani 2024^85^ |
| COVID | Lithium | Mood stabilizer | Mania, Manic-depressive illness, Recurrent depression, Aggressive or self-mutilating behaviour, Bipolar | No | Zeidan 2024^2^ |
| COVID | Everolimus | mTOR (mechanistic Target of Rapamycin) inhibitor | Neuroendocrine tumours of pancreatic origin, Neuroendocrine tumours of gastrointestinal or lung origin, Renal cell carcinoma | Yes | Barghash 2024,^4^ Godde 2024,^70^ Ullah 2024,^90^ Voloudakis 2025,^81^ Xu 2024^69^ |
| COVID | Sirolimus | mTOR (mechanistic Target of Rapamycin) inhibitor | Organ rejection, Sporadic lymphangioleiomyomatosis | No | Bogacheva 2024,^78^ Godde 2024,^70^ Ullah 2024,^90^ Xu 2024^69^ |
| COVID | Dornase alfa | Mucolytic enzyme | Cystic fibrosis | No | Kumawat 2024^54^ |
| COVID | Aprepitant | Neurokinin-1 receptor antagonist | Prevent nausea and vomiting | No | Godde 2024^70^ |
| COVID | Timolol maleate | Non-selective beta-adrenergic blocker | Elevated intra-ocular pressure, Ocular hypertension, Secondary glaucoma | No | Alzahrani 2024^52^ |
| COVID | Celecoxib | Nonsteroidal anti-inflammatory drug (NSAID) | Osteoarthritis, Rheumatoid arthritis, Ankylosing spondylitis | No | Martins 2024^63^ |
| COVID | Naproxen | Nonsteroidal anti-inflammatory drug (NSAID) | Rheumatoid arthritis, Osteoarthrosis, Ankylosing spondylitis, Gout, Acute musculoskeletal disorders, Dysmenorrhoea | No | Martins 2024^63^ |
| COVID | Aspirin | Nonsteroidal anti-inflammatory drug (NSAID), antiplatelet | Analgesic, Antipyretic, Anti-inflammatory actions, Secondary prevention of thrombotic cerebrovascular or cardiovascular disease, Anti-thrombotic action | No | Jaimes-Castelan 2024^48^ |
| COVID, MERS, SARS | Ribavirin | Nucleoside antiviral (RNA synthesis inhibitor) | In combination with other drugs for: Chronic hepatitis C | No | Barghash 2024,^4^ Boulon 2024,^49^ Chan 2024,^91^ Chavda 2024,^60^ Jaimes-Castelan 2024,^48^ Kumawat 2024,^54^ Mia 2024^50^ |
| COVID | Zidovudine | Nucleoside reverse transcriptase inhibitor | Human Immunodeficiency Virus | No | Alzahrani 2024,^52^ Gao 2024^71^ |
| COVID | Tenofovir | Nucleotide reverse transcriptase inhibitor | Chronic hepatitis B | No | Gao 2024,^71^ Kumawat 2024,^54^ Singh 2024,^64^ Waseem 2024^73^ |
| COVID | Cobicistat | Pharmacokinetic enhancer | In combination with other drugs for: Human Immunodeficiency Virus | No | Gallucci 2024,^92^ Pereira 2024^68^ |
| COVID, MERS, SARS | Cobicistat/darunavir | Pharmacokinetic enhancer + protease inhibitor | In combination with other drugs for: Human Immunodeficiency Virus | No | Barghash 2024^4^ |
| COVID | Duvelisib | Phosphoinositide 3-kinase inhibitor | Chronic lymphocytic leukaemia, Follicular lymphoma | Yes | Bogacheva 2024^78^ |
| COVID | Temoporfin | Photosensitizer | Head and neck squamous cell carcinoma | Yes | Mendonca 2024^93^ |
| COVID | Verteporfin | Photosensitizer | Exudative age-related macular degeneration, Subfoveal choroidal neovascularisation secondary to pathological myopia | No | Mendonca 2024^93^ |
| COVID | Cannabidiol | Phytocannabinoid | Seizures associated with tuberous sclerosis complex, Lennox-Gastaut syndrome, Multiple sclerosis | No | Nazir 2024^51^ |
| COVID | Rucaparib | Poly (ADP-ribose) polymerase (PARP) inhibitor | Epithelial ovarian, Fallopian tube, or Primary peritoneal cancer | Yes | Papp 2024^89^ |
| COVID | Amiloride | Potassium-sparing diuretic | Oedema, Hypertension | No | Brun 2025^53^ |
| COVID | Aprotinin | Protease inhibitor | Reduce blood loss, Blood transfusion | No | Padin 2024^94^ |
| COVID | Atazanavir | Protease inhibitor | In combination with ritonavir for: Human Immunodeficiency Virus | No | Metwaly 2024^56^ |
| COVID | Darunavir | Protease inhibitor | Human Immunodeficiency Virus | No | Chavda 2024,^60^ Jaimes-Castelan 2024,^48^ Marinho 2025,^72^ Martins 2024,^63^ Metwaly 2024,^56^ Paroczai 2024,^86^ Pereira 2024^68^ |
| COVID | Lopinavir | Protease inhibitor | In combination with ritonavir (fixed dose combination) for: Human Immunodeficiency Virus | No | Chakraborty 2024,^65^ Chavda 2024,^60^ Enyeji 2024,^57^ Handa 2024,^95^ Hongyu 2024,^96^ Jaimes-Castelan 2024,^48^ Marinho 2025,^72^ Mawazi 2024,^67^ Metwaly 2024,^56^ Mia 2024,^50^ Paroczai 2024,^86^ Singh 2024,^64^ Barghash 2024^4^ |
| COVID | Lopinavir/Ritonavir | Protease inhibitor | Human Immunodeficiency Virus | No | Enyeji 2024,^57^ Hongyu 2024,^96^ Mawazi 2024,^67^ Mia 2024^50^ |
| COVID, MERS, SARS | Lopinavir/Ritonavir + ribavirin | Protease inhibitor + Nucleoside antiviral (RNA synthesis inhibitor) | Human Immunodeficiency Virus \| In combination with other medicinal products for: Chronic hepatitis C | No | Enyeji 2024,^57^ Hongyu 2024,^96^ Mawazi 2024,^67^ Mia 2025^50^ |
| COVID | Carfilzomib | Proteasome inhibitor | In combination with other drugs for: Multiple myeloma | Yes | Metwaly 2024,^56^ Low 2025,^84^ Rahmani 2024^85^ |
| COVID | Fluoxetine | Selective serotonin reuptake inhibitor | Major depressive disorders, Obsessive-compulsive disorder, Bulimia nervosa | No | Kumawat 2024^54^ |
| COVID | Fluvoxamine | Selective serotonin reuptake inhibitor | Major depressive episode, obsessive compulsive disorder | No | Godde 2024,^70^ Jaimes-Castelan 2024,^48^ Prasanth 2024,^97^ Singh 2024,^64^ Wannigama 2024^98^ |
| COVID | Cefiderocol | Siderophore cephalosporin antibiotic | Gram-negative Infections | No | Ahmad 2024^74^ |
| COVID | Fostamatinib | Spleen tyrosine kinase inhibitor | Chronic immune thrombocytopenia | No | Bakshi 2025^99^ |
| COVID | Gliclazide | Sulfonylurea | Non-insulin-dependent diabetes | No | Yip 2025^75^ |
| COVID | Dexamethasone | Systemic corticosteroid | Non-infectious inflammatory conditions affecting the anterior segment of the eye, Cerebral oedema, Asthma, Skin diseases, Systemic lupus erythematodes, Systemic vasculitides, Rheumatoid arthritis, Still's disease, Idiopathic thrombocytopenic purpura, Tuberculous meningitis, Neoplastic diseases, Emesis induced by cytostatics, Emetogenic chemotherapy, Multiple myeloma, Acute lymphocytic leukaemia, Acute lymphoblastic leukaemia, General antiemetic treatment | No | Chavda 2024,^60^ Jaimes-Castelan 2024,^48^ Martins 2024,^63^ Okeowo 2024,^88^ Papp 2024,^89^ Paroczai 2024^86^ |
| COVID | Methylprednisolone | Systemic corticosteroid | Rheumatoid arthritis, Lupus erythematosus, Stevens-Johnson syndrome, Bronchial asthma, Drug hypersensitivity reactions, Angioneurotic oedema, Ulcerative colitis, Crohn's disease, Tuberculosis, Aspiration of gastric contents, Tuberculosis meningitis, Osteo-arthritis with an inflammatory component, Synovitis not associated with infection, Epicondylitis, Tenosynovitis, Plantar fasciitis, Bursitis, Keloids, Localized lichen planus, Localized lichen simplex, Granuloma annulare, Alopecia areata | No | Enyeji 2024,^57^ Jaimes-Castelan 2024,^48^ Martins 2024^63^ |
| COVID | Doxycycline | Tetracycline antibiotic | Respiratory tract infections, Urinary tract infections, sexually transmitted infections, Skin infections, Eye infections, Rickettsial infections, Rosacea, cholera, Bubonic plague, Louse and tick-borne relapsing fever, Tularaemia glanders, Melioidosis, Chloroquine-resistant falciparum malaria, Acute intestinal amoebiasis | No | Enyeji 2024^57^ |
| COVID | Minocycline | Tetracycline antibiotic | Acne | No | Chakraborty 2024^65^ |
| COVID | Etopophos | Topoisomerase II inhibitor | In combination with other drugs for: Testicular cancer, small cell lung cancer, Hodgkin's lymphoma, non-Hodgkin’s lymphoma, Acute myeloid leukaemia, Gestational trophoblastic neoplasia, Ovarian cancer | Yes | Alzahrani 2024^52^ |
| COVID | Entrectinib | Tyrosine kinase inhibitor | Solid tumours, non-small cell lung cancer | Yes | Ahmad 2024,^74^ Xu 2024^69^ |
| COVID | Imatinib | Tyrosine kinase inhibitor | Chronic myelogenous leukaemia, Acute lymphoblastic leukaemia, Myelodysplastic/myeloproliferative diseases, Hypereosinophilic syndrome, Gastrointestinal stromal tumours, Dermatofibrosarcoma protuberans | Yes | Ghavimehr 2024,^100^ Jaimes-Castelan 2024,^48^ Low 2025,^84^ Rahmani 2024^85^ |
| COVID | Nilotinib | Tyrosine kinase inhibitor | Chronic myelogenous leukaemia | Yes | Ghavimehr 2024,^100^ Malar 2024,^101^ Xu 2024^69^ |
| COVID | Vandetanib | Tyrosine kinase inhibitor | Thyroid cancer | Yes | Puhl 2023^102^ |
| COVID | Bevacizumab | Vascular Endothelial Growth Factor inhibitor | In combination with other drugs for: Carcinoma of the colon or rectum, Breast cancer, non-small cell lung cancer, Renal cell cancer, Epithelial ovarian, Fallopian tube, Primary peritoneal cancer, Cervical cancer | Yes | Jaimes-Castelan 2024^48^ |
| COVID | Tivozanib | Vascular Endothelial Growth Factor inhibitor | Renal cell carcinoma | Yes | Alzahrani 2024^52^ |
| COVID | Vincristine sulfate | Vinca alkaloid | Leukaemias, Lymphoma, Multiple myeloma, Solid tumours, Idiopathic thrombocytopenic purpura | Yes | Alzahrani 2024^52^ |
| COVID | Oseltamivir |  | Influenza | No | Gao 2024,^71^ Jaimes-Castelan 2024,^48^ Marinho 2025,^72^ Mia 2024,^50^ Waseem 2024,^73^ Barghash 2024^4^ |
| COVID | Ritonavir |  | Human Immunodeficiency Virus | No | Boulon 2024,^49^ Chavda 2024,^60^ Enyeji 2024,^57^ Gallucci 2024,^92^ Godde 2024,^70^ Hongyu 2024,^96^ Jaimes-Castelan 2024,^48^ Marinho 2025,^72^ Metwaly 2024,^56^ Mia 2024,^50^ Paroczai 2024,^86^ Singh 2024,^64^ Uzuner 2024^103^ |

**Supplementary Table 6 – Interventional clinical trials identified for influenza**

| **Medicines** | **Therapeutic Class** | **UK/EU Approved Indication** | **Cancer Y/N** | **Trial ID** | **Trial Phase** |
| --- | --- | --- | --- | --- | --- |
| Molnupiravir | Antiviral (RNA polymerase inhibitor) | COVID-19 | No | NCT05648448 ^104^ | Phase II |
| Dexamethasone sodium phosphate | Corticosteroid | Autoimmune disorders, rheumatology, oncology | No | NCT04366115 ^132^ | Phase I |
| Sirolimus + Oseltamivir | mTOR (mechanistic Target of Rapamycin) inhibitor + Antiviral (Neuraminidase inhibitor) | Sporadic lymphangioleiomyomatosis, organ rejection \| Influenza | No | NCT03901001 ^105^ | Phase III |
| N-acetylcysteine + Oseltamivir | Mucolytic agent + Antiviral (Neuraminidase inhibitor) | Respiratory tract diseases, Influenza | No | NCT03900988 ^106^ | Phase III |

**Supplementary Table 7 –** **Interventional clinical trials identified for COVID-19**

| **Medicines** | **Therapeutic Class (A-Z)** | **UK/EU Approved Indication** | **Cancer Y/N** | **Trial ID** | **Trial Phase** |
| --- | --- | --- | --- | --- | --- |
| Captopril | Angiotensin-Converting Enzyme (ACE) Inhibitors | Hypertension, Myocardial Infarction | No | NCT04345406 ^107^ | Phase III |
| Colchicine | Antigout agent | Acute gout | No | NCT04381936 ^108^ | Phase III |
| Hydroxychloroquine | Antimalarial and immunomodulatory agent | Rheumatoid arthritis, discoid and systemic lupus erythematosus | No | NCT05041907 ^109^ | Phase II |
| Hydroxychloroquine sulfate | Antimalarial and immunomodulatory agent | Rheumatoid arthritis, discoid and systemic lupus erythematosus | No | NCT05113810 ^110^ | Phase II |
| Hydroxychloroquine | Antimalarial and immunomodulatory agent | Rheumatoid arthritis, discoid and systemic lupus erythematosus | No | NCT04381936 ^108^ | Phase III |
| Hydroxychloroquine | Antimalarial and immunomodulatory agent | Rheumatoid arthritis, discoid and systemic lupus erythematosus | No | NCT04315948 ^111^ | Phase III |
| Hydroxychloroquine + Azithromycin | Antimalarial and immunomodulatory agent + Macrolide antibiotic | Rheumatoid arthritis, discoid and systemic lupus erythematosus\| Bacterial infections | No | NCT04334512 ^112^ | Phase II |
| Ivermectin | Antiparasitic | Intestinal strongyloidiasis, microfilaraemia, human sarcoptic scabies | No | NCT05041907 ^109^ | Phase II |
| Ivermectin | Antiparasitic | Intestinal strongyloidiasis, microfilaraemia, human sarcoptic scabies | No | NCT04351347 ^113^ | Phase II \|Phase III |
| Ivermectin | Antiparasitic | Intestinal strongyloidiasis, microfilaraemia, human sarcoptic scabies | No | NCT04703608 ^114^ | Phase III |
| Ivermectin | Antiparasitic | Intestinal strongyloidiasis, microfilaraemia, human sarcoptic scabies | No | NCT04885530 ^115^ | Phase III |
| Ivermectin | Antiparasitic | Intestinal strongyloidiasis, microfilaraemia, human sarcoptic scabies | No | NCT05040724 ^116^ | Phase III |
| Ivermectin | Antiparasitic | Intestinal strongyloidiasis, microfilaraemia, human sarcoptic scabies | No | NCT05736861 ^117^ | Phase III |
| Ivermectin | Antiparasitic | Intestinal strongyloidiasis, microfilaraemia, human sarcoptic scabies | No | NCT05894538 ^118^ | Phase III |
| Ivermectin + Doxycycline | Antiparasitic + Tetracycline antibiotic | Intestinal strongyloidiasis, microfilaraemia, human sarcoptic scabies\| Papulopustular lesions | No | NCT04482686 ^119^ | Phase I |
| Remdesivir + Baricitinib | Antiviral (RNA polymerase inhibitor) + Janus kinase (JAK) inhibitor | COVID-19 \| Rheumatoid arthritis, Atopic dermatitis, Alopecia areata, Juvenile idiopathic arthritis | No | NCT04321993 ^120^ | Phase II |
| Remdesivir + Dexamethasone + Apremilast | Antiviral (RNA polymerase inhibitor) + Corticosteroid + Phosphodiesterase inhibitors | COVID-19 \| Autoimmune disorders, rheumatology, oncology \| Psoriatic arthritis, psoriasis, oral ulcers | No | NCT04488081 ^121^ | Phase II |
| Remdesivir + Dexamethasone + Celecoxib + Famotidine | Antiviral (RNA polymerase inhibitor) + Corticosteroid + Nonsteroidal anti-inflammatory drug (NSAID) + Histamine H2-receptor antagonists | COVID-19 \| Autoimmune disorders, rheumatology, oncology \| Osteoarthritis, rheumatoid arthritis, ankylosing spondylitis \| Zollinger-Ellison syndrome, gastric ulcer, mild reflux oesophagitis | No | NCT04488081^121^ | Phase II |
| Remdesivir + Dexamethasone + ciclosporin | Antiviral (RNA polymerase inhibitor) + Corticosteroid + Immunosuppressants | COVID-19 \| Autoimmune disorders, rheumatology, oncology \| Endogenous uveitis, Rheumatoid arthritis, Psoriasis, Atopic dermatitis, Transplantation indications | No | NCT04488081^121^ | Phase II |
| Remdesivir + Dexamethasone + Dornase alfa | Antiviral (RNA polymerase inhibitor) + Corticosteroid + Recombinant human deoxyribonuclease I | COVID-19 \| Autoimmune disorders, rheumatology, oncology \| Cystic fibrosis | No | NCT04488081^121^ | Phase II |
| Remdesivir + Dexamethasone + Icatibant | Antiviral (RNA polymerase inhibitor) + Corticosteroid + Bradykinin B2 receptor antagonists | COVID-19 \| Autoimmune disorders, rheumatology, oncology \| Hereditary angioedema | No | NCT04488081^121^ | Phase II |
| Remdesivir + Imatinib Mesylate + Dexamethasone | Antiviral (RNA polymerase inhibitor) + Tyrosine-kinase inhibitors + Corticosteroid | COVID-19 \| Chronic myeloid leukaemia, gastrointestinal stromal tumours \| Autoimmune disorders, rheumatology, oncology | No | NCT04488081^121^ | Phase II |
| Amantadine Hydrochloride | Antiviral and antiparkinsonian | Parkinson's disease | No | NCT04854759 ^122^ | Phase III |
| Metformin | Biguanide | Type 2 diabetes mellitus | No | NCT05041907 ^109^ | Phase II |
| Metformin + Ivermectin | Biguanide + Antiparasitic | Type 2 diabetes mellitus \| Intestinal strongyloidiasis, microfilaraemia, human sarcoptic scabies | No | NCT04510194 ^123^ | Phase III |
| Metformin + Fluvoxamine | Biguanide + Selective Serotonin Reuptake Inhibitors (SSRIs) | Type 2 diabetes mellitus \| Major depressive episode, obsessive compulsive disorder | No | NCT04510194^123^ | Phase III |
| Budesonide | Corticosteroid | Seasonal allergic rhinitis | No | NCT04727424 ^124^ | Phase III |
| Fluticasone | Corticosteroid | Allergic rhinitis | No | NCT04885530 ^115^ | Phase III |
| Interferon beta 1a | Immunomodulator | Multiple sclerosis | No | NCT04518410 ^125^ | Phase II \|Phase III |
| Interferon beta-1b + Ribavirin | Immunomodulator + Nucleoside antiviral (RNA synthesis inhibitor) | Multiple sclerosis\| Chronic hepatitis C | No | NCT04494399 ^126^ | Phase II |
| Dimethyl fumarate | Immunomodulators | Multiple sclerosis | No | NCT04381936 ^108^ | Phase III |
| Anakinra | Interleukin-1 receptor antagonist | Rheumatoid Arthritis, Periodic fever syndromes, Still's Disease | No | NCT04381936^108^ | Phase III |
| Baricitinib | Janus kinase (JAK) inhibitor | Rheumatoid arthritis, Atopic dermatitis, Alopecia areata, Juvenile idiopathic arthritis | No | NCT04321993 ^120^ | Phase II |
| Baricitinib | Janus kinase (JAK) inhibitor | Rheumatoid arthritis, Atopic dermatitis, Alopecia areata, Juvenile idiopathic arthritis | No | NCT04381936 ^108^ | Phase III |
| Montelukast | Leukotriene receptor antagonist | Asthma | No | NCT04389411 ^127^ | Phase II \|Phase III |
| Montelukast | Leukotriene receptor antagonist | Asthma | No | NCT04885530 ^115^ | Phase III |
| Azithromycin | Macrolide antibiotic | Bacterial infections | No | NCT04381936 ^108^ | Phase III |
| Omalizumab | Monoclonal antibody | Allergic asthma, Chronic rhinosinusitis with nasal polyps | No | NCT04720612 ^128^ | Phase II |
| Tocilizumab | Monoclonal antibody | Rheumatoid arthritis, juvenile idiopathic arthritis, Giant Cell Arteritis | No | NCT04321993 ^120^ | Phase II |
| Aspirin | Nonsteroidal anti-inflammatory drug (NSAID), antiplatelet | Thrombotic cerebrovascular or cardiovascular disease, inflammatory conditions | No | NCT04381936 ^108^ | Phase III |
| Emtricitabine/Tenofovir Disoproxil Fumarate | Nucleoside Reverse Transcriptase Inhibitor (NRTI) | Human immunodeficiency virus (HIV-1) | No | NCT04890626 ^129^ | Phase III |
| Lopinavir/ritonavir | Protease inhibitor | Human immunodeficiency virus (HIV-1) | No | NCT05925140 ^130^ | Phase I |
| Lopinavir/ritonavir | Protease inhibitor | human immunodeficiency virus (HIV-1) | No | NCT04315948 ^111^ | Phase III |
| Lopinavir/ritonavir | Protease inhibitor | Human immunodeficiency virus (HIV-1) | No | NCT04381936 ^108^ | Phase III |
| Lopinavir/ritonavir + Interferon Beta-1A | Protease inhibitor + Immunomodulator | Human immunodeficiency virus (HIV-1) \| Multiple sclerosis | No | NCT04315948 ^111^ | Phase III |
| Fluoxetine | Selective serotonin reuptake inhibitor | Major depressive episodes, obsessive-compulsive disorder | No | NCT05041907 ^109^ | Phase II |
| Fluvoxamine | Selective serotonin reuptake inhibitor | Major depressive episodes, obsessive-compulsive disorder | No | NCT04885530^115^ | Phase III |
| Fluvoxamine | Selective serotonin reuptake inhibitor | Major depressive episode, obsessive compulsive disorder | No | NCT05894564 ^131^ | Phase III |
| Empagliflozin | Sodium-Glucose Co-Transporter 2 (SGLT2) Inhibitor | Type 2 diabetes mellitus, Heart failure, chronic kidney disease | No | NCT04381936 ^108^ | Phase III |
| Dexamethasone sodium phosphate | Systemic corticosteroid | Non-infectious inflammatory conditions affecting the anterior segment of the eye, Cerebral oedema, Asthma, Skin diseases, Systemic lupus erythematodes, Systemic vasculitides, Rheumatoid arthritis, Still's disease, Idiopathic thrombocytopenic purpura | No | NCT04366115^132^ | Phase I |
| Dexamethasone | Systemic corticosteroid | Non-infectious inflammatory conditions affecting the anterior segment of the eye, Cerebral oedema, Asthma, Skin diseases, Systemic lupus erythematodes, Systemic vasculitides, Rheumatoid arthritis, Still's disease, Idiopathic thrombocytopenic purpura | No | NCT04381936 ^108^ | Phase III |
| Infliximab | Tumour necrosis factor (TNF) inhibitor | Rheumatoid arthritis, Crohn's disease, Ulcerative colitis, Ankylosing spondylitis, Psoriatic arthritis, Psoriasis | No | NCT04330690 ^133^ | Phase III |
| Imatinib Mesylate | Tyrosine kinase inhibitor | Chronic myeloid leukaemia, gastrointestinal stromal tumours | Yes | NCT04488081 ^121^ | Phase II |
| Imatinib mesilate + Infliximab | Tyrosine kinase inhibitor + Tumour necrosis factor (TNF) inhibitor | Chronic myeloid leukaemia, gastrointestinal stromal tumours \| Rheumatoid arthritis, Crohn's disease, Ulcerative colitis, Ankylosing spondylitis, Psoriatic arthritis, Psoriasis | Yes | NCT05220280 ^134^ | Phase IV |
